# Persistent Hypercoagulability and Further Characterization of Microclot Complexes in Long COVID

**DOI:** 10.64898/2026.08.20.26360874

**Authors:** Massimo Nunes, Carlos M Pereira Guerreiro, Jan H Pretorius, Chantelle Venter, Alain R. Thierry, Burtram C Fielding, Douglas B Kell, Etheresia Pretorius

## Abstract

**Background:** Growing evidence suggests persistent thrombotic endothelial damage (together with elevated (fibrinaloid) microclot complexes (FMCs)) and immune dysfunction in the pathophysiology of Long COVID. Recently we proposed that there are different FMC phenotypes. Here we seek to determine the nature of these FMCs and aggregates in platelet-poor plasma (PPP) by using different markers, as well as thromboelastography (TEG^®^) to assess for hypercoagulability of samples.

**Material and Methods:** Whole-blood and PPP from control (n=19) and Long COVID (n=20) participants were assessed by thromboelastography. FMCs were quantified by imaging flow cytometry of Thioflavin-T (ThT)-stained PPP, 10X diluted PPP, and resuspended PPP pellets. The resuspended pellets were separately stained with a CD62P-PE antibody or Hoechst 33342 to label aggregates and FMCs containing amyloid, platelet, and nuclear material. ThT and CellMask™ Red were co-stained for confocal microscopy. ThT and myeloperoxidase (MPO), and ThT, Congo Red, and Hoechst were co-stained for fluorescence and polarized microscopy. Whole-blood smears were imaged by scanning electron microscopy (SEM).

**Results:** Long COVID samples showed pronounced hypercoagulability in both whole blood and PPP, with shortened R, K and TMRTG and elevated α-angle and MRTG, but unchanged MA and TTG, indicating altered clotting kinetics. Persistence of this phenotype in PPP implicates soluble plasma constituents. ThT-positive FMCs were significantly increased in Long COVID across undiluted, diluted, and resuspended pellet samples; counts were processing-sensitive and a substantial ThT-positive population remained in the supernatant after centrifugation, indicating heterogeneity in density. Across probes, leukocyte material was the most abundant, then platelet material, and ThT-positive FMCs were the least abundant, with the three populations exhibiting unique morphology and occupying distinct size domains. Platelet-derived material was significantly elevated in Long COVID, whereas nuclear material was not. Co-stained samples subject to confocal, fluorescence, and polarized microscopy imaging showed that FMCs are heterogeneous, including events positive for ThT, CellMask™, Hoechst, MPO, and Congo Red, and also a distinct subset of membrane-free, ThT-only events.

**Conclusion:** In this Long COVID cohort, plasma is characterised by hypercoagulability and an increased burden of ThT-positive FMCs that are numerically minor relative to, and morphologically distinct from, aggregates and amyloidogenic FMCs marked with platelet- and leukocyte-derived material. The increased burden of platelet debris in PPP is likely indicative of persistent platelet activity. The existence of membrane-free, ThT-only FMCs, in addition to FMCs associated with cellular material, confirms an amyloid-dominated FMC population. Furthermore, positive Congo Red signal further confirms the amyloid nature of FMCs in PPP.

**Graphical Abstract (created with Biorender.com):** 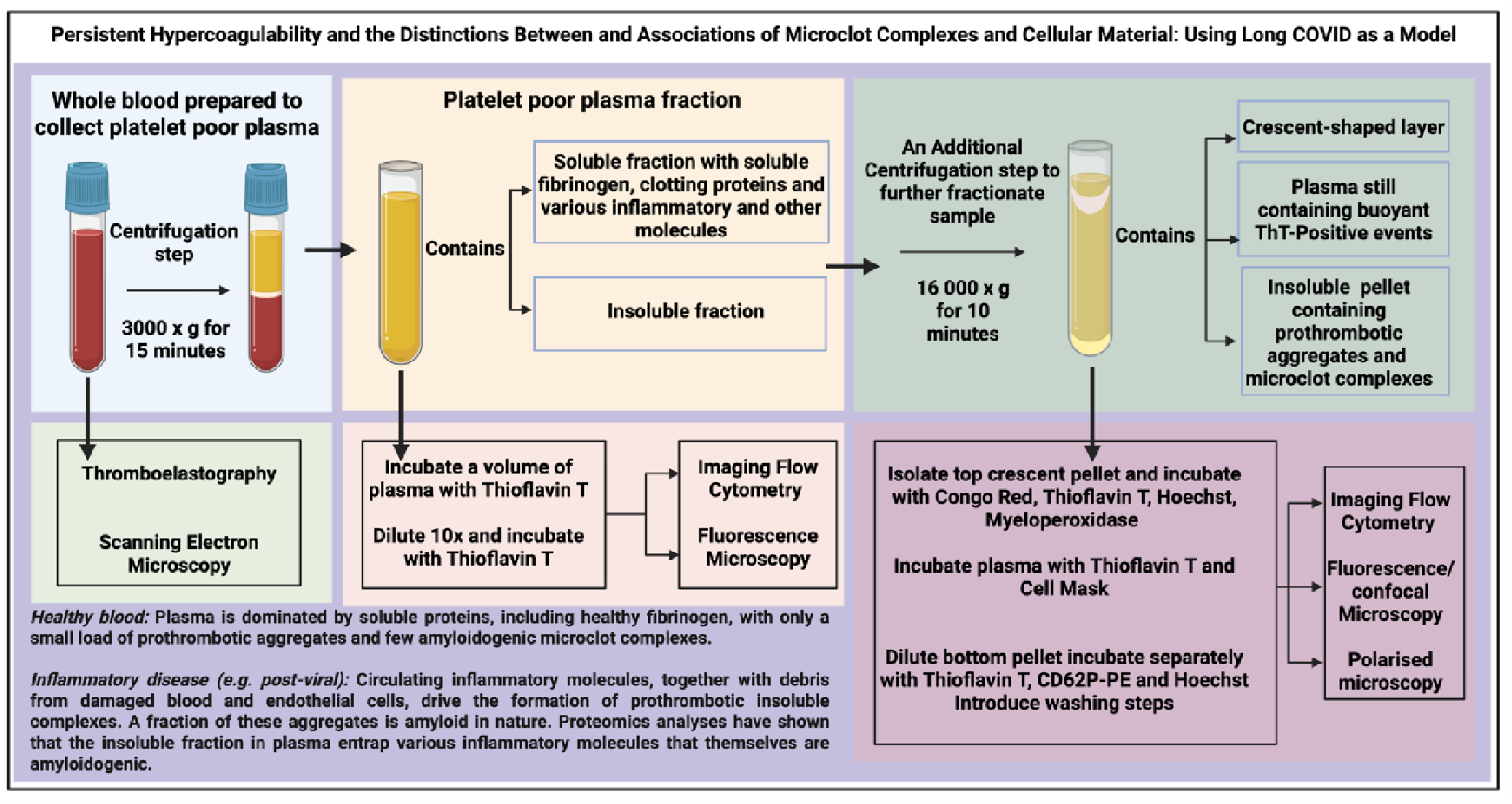

## Introduction

Since the acute phase of the COVID-19 pandemic, a subset of individuals infected with SARS-CoV-2 have developed persistent, systemic symptoms lasting months or longer after the resolution of the initial infection. By 2024, it was estimated that hundreds of millions of individuals worldwide have been affected by this condition [1], commonly termed Long COVID or post-acute sequelae of SARS-CoV-2 infection (PASC). Long COVID is characterised by heterogeneous clinical manifestations including chronic fatigue, post-exertional malaise (PEM), cognitive dysfunction, orthostatic intolerance, respiratory issues, musculoskeletal pain, gastrointestinal disturbance, sleep disruption, and more [2–4]. Despite advances in recent years [5], the underlying pathophysiological mechanisms of Long COVID still require further elucidation to establish clinical biomarkers and effective treatment regimens.

Accumulating evidence implicates dysfunction of the vascular and coagulation system as a consistent feature of Long COVID [6–12], among other pathomechanisms [5]. This includes dysregulated platelets [13], abnormal coagulation factor levels [14], increased clotting kinetics [15], increases in D-dimer [11, 16], impairments in fibrinolysis [17], and elevations in neutrophil extracellular traps (NETs) [18, 19] and (fibrinaloid) microclot complexes (FMCs) [8, 20–23].

As recently formalised [24, 25], FMCs, which range from submicron to 600µm^2^ (and larger in some cases) in area [21, 26], are a class of circulating insoluble complexes that demonstrate resistance to fibrinolysis [27, 28], exhibit an amyloid phenotype as determined by thioflavin T (ThT) positivity [27–30], entrap a variety of plasma and cellular proteins [31–34], and have been shown to associate with NETs [21, 35]. It has also been shown that FMCs can be induced in a purified fibrinogen model, in response to inflammagens originating from viruses or bacteria [25, 27–30, 36, 37].

FMCs can also arise when fibrin(ogen) and amyloidogenic inflammatory proteins co-aggregate on damaged, procoagulant membrane surfaces, distinct from canonical thrombin-driven fibrin clots [24]. Rather than a single uniform species, FMCs exhibit several content-based phenotypes: (i) cell-derived debris: large fragments of dying or apoptotic cells retaining phosphatidylserine (PS)-exposed, β-sheet-rich membrane patches (e.g. hyperactivated platelets); (ii) subcellular vesicles and microparticles (∼0.1-1µm) bearing membrane proteins and PS that bind fibrinogen and coagulation factors; (iii) nucleoprotein immune complexes such as NETs; and (iv) plasma-protein aggregates in which fibrin(ogen) associates with amyloidogenic molecules such as serum amyloid A, von Willebrand factor, or viral proteins. This framework positions FMCs as heterogeneous complexes whose composition varies with the underlying inflammatory trigger. Platelet-poor plasma (PPP) thus contains a heterogeneous mixture of (decaying) cell-derived material originating from leukocytes, platelets, erythrocytes and, to a lesser extent, endothelial cells [38]. Cellular debris in blood is itself proinflammatory [38].

Although the role of amyloid-β (Aβ) peptides in the context of FMCs has not yet been explored, there are indications that platelets might significantly contribute to their amyloidogenic properties by the release of Aβ [39, 40]. Platelets have been shown to store pre-formed amyloid-β (Aβ) peptides, including Aβ_₁- ₄₂_, within α-granules, which are rapidly released upon activation by thrombin, lipopolysaccharide (LPS), collagen, and arachidonic acid [41, 42]. Inhibition of β secretase or cyclooxygenase enzymes did not reduce the load of Aβ peptides, indicating that platelet activation triggers the release of pre-formed Aβ rather than requiring acute amyloid precursor protein (APP) processing. This suggests that activated platelets in inflammatory and thrombotic environments may be a source of extracellular Aβ, and that platelet-derived Aβ has the potential to contribute to the both the amyloidogenicity and persistence of FMCs (specifically the ones originating from hyperactivated platelets), by stabilizing the amyloid architecture of these structures. The two concepts, cell-derived debris and FMCs, as well as cell-derived microparticles [43], therefore lie on a continuum rather than forming separate categories, ranging from non-amyloid (but prothrombotic) complexes to amyloid-bearing complexes.

FMCs have been shown to be elevated in Long COVID [8, 20, 22, 24, 30, 32] and myalgic encephalomyelitis/chronic fatigue syndrome (ME/CFS) [44], as well as other inflammatory conditions such as diabetes mellitus type II [45] and sepsis [46]. The persistence of and elevations in FMCs, along with platelet dysregulation and increased clotting propensities, have been hypothesized to contribute to endothelial and microvascular dysfunction, impaired microvascular perfusion, tissue hypoxia, and ultimately symptom manifestation [47]. The thrombi removed following ischaemic stroke have also been shown to be amyloid in character [48, 49].

There is therefore a need to quantify ThT-positive (amyloid-bearing) aggregates alongside the broader pool of cell-derived material, in order to identify, classify and interpret these events and to establish what fraction of the prothrombotic debris in PPP has acquired amyloid character. The present study aimed to measure coagulability, FMC concentrations across various PPP sample types, including undiluted PPP, diluted PPP, and resuspended PPP pellets in control and Long COVID cohorts. Thromboelastography (TEG^®^) [50] was used to assess global coagulation kinetics and clot strength and load. Imaging flow cytometry, confocal microscopy, and scanning electron microscopy was also used to quantify, visualise, and characterise FMC subtypes. This study provides updated insights into the coagulation profile of individuals with Long COVID, the concentrations and phenotypes of FMCs, and novel observations regarding cell-derived material and its relationship to FMCs in plasma samples.

## Materials and Methods

### Ethical clearance and sample cohorts and collection

See supplementary material for details regarding ethical clearance, and sample cohort and collection. 19 healthy controls (14 females; 5 males) and 20 Long COVID (13 females; 7 males) were recruited for this study (Figure S1 and Table S1).

### Thromboelastography^®^ Analysis of Whole-Blood and Platelet-Poor Plasma

The viscoelastic properties of whole-blood and PPP was assessed using the Thrombelastograph^®^ (TEG^®^) 5000 Hemostasis Analyzer (Haemoscope Corp). These analyses were performed on fresh samples on the day of blood collection. Because coagulation was being assessed, samples were not treated with platelet inhibitors (see Supplementary Material).

### Imaging Flow Cytometry

There are a few differences between measuring FMCs and the traditional coagulation cascade in blood samples. FMCs are formed and circulate in blood, and are then obtained in anticoagulant tubes during blood collection. Hence, to measure FMCs there is no need to induce coagulation in blood samples once collected; this is in contrast to typical coagulation assessments, such as TEG^®^, where clot induction is required. This, together with the size range of FMCs; from submicron to 600 µm² in area (and occasionally larger, though rarely); enables their detection and quantification by microscopy or by more quantitative techniques such as flow cytometry.

Five staining conditions were assessed by imaging flow cytometry, each performed as a separate experiment: (1) undiluted PPP stained with ThT (Sigma-Aldrich, St. Louis, MO, USA); (2) PPP diluted 10X in PBS and stained with ThT; and the resuspended PPP pellet stained separately with (3) ThT, (4) or CD62P-PE antibody (IM1759U, Beckman Coulter, Brea, CA, USA), or (5) Hoechst 33342 (Invitrogen, Thermo Fisher Scientific). Frozen PPP samples were thawed and allowed to equilibrate to room temperature before staining. As platelets and leukocytes are largely absent from PPP, CD62P and Hoechst staining primarily identify platelet- and nuclear-derived debris rather than whole cells. For the undiluted condition, a ThT exposure concentration of 15 µM was used. For the diluted condition, 4.7 μL PPP was first diluted with 42.3 μL PBS and then stained with 3 μL ThT (final exposure concentration of 15 µM).

To obtain the remaining three conditions, PPP was centrifuged at 16,000 x g for 10 minutes at room temperature, which reproducibly yielded two distinct insoluble fractions. The first was a dense pellet at the base of the tube. The supernatant was collected and retained for separate ThT staining and imaging, and the pellet was resuspended in 400 μL PBS. The sample was then stained with either ThT, or CD62P-PE antibody, or Hoechst 33342, and then washed twice with PBS at 3000xg. The resuspended pellet was exposed to ThT or Hoechst at an exposure concentration of 15 µM and 2 µM, respectively, or to the CD62P-PE antibody, by adding 4 μL of the stock solution to 46 μL of sample. All staining was performed in the dark for 30 minutes before data acquisition.

The second fraction was a crescent-shaped deposit adhering near to the top of the side of the tube, approximately opposite the basal pellet (as depicted in the graphical abstract) and consistent with a lipid fraction containing some FMCs. To be able to study this “crescent pellet”, PPP samples were centrifuged at 16,000 x g for 10 minutes at room temperature and then another 10 minutes at 0°C. Unfortunately, due to the nature of this pellet, it could only be partly retrieved, and it was then resuspended in 400µL of PBS.

Data were acquired on a Cytek^®^ Amnis^®^ FlowSight^®^ imaging flow cytometer (Luminex) running INSPIRE™ (Amnis^®^) software (version 200.0.336.0). The instrument was calibrated according to the manufacturer’s instructions, and each sample was vortexed before use. The 405 nm and side-scatter lasers were used for ThT- and Hoechst-stained samples, and the 488 nm and side-scatter lasers for samples stained with the CD62P-PE antibody. Images were captured with a 20x objective. ThT- and Hoechst-positive events were recorded in channel 7 (emission 437-485 nm), CD62P-PE-positive events in channel 3 (emission 560-595 nm), and brightfield images in channel 1. All samples were acquired at a flow rate of 0.043 µL/s for 5 minutes. Details of the acquisition and analysis gates and the masking strategy are provided in the Supplementary Material Figure S2 to S4).

For each sample, the INSPIRE™ software returned the total event count, the percentage of gated events, and the event count within each of six area bins: 0–1, 1–10, 10–30, 30–100, 100–400, and 400–3000 µm². Because the analysis gate spans events ranging from submicron to several hundred microns in area, the mean area across all events is not a meaningful summary of event size; size was therefore characterised by the distribution of events across these bins.

### Confocal laser scanning microscopy

In order to assess the association of FMCs with cellular material, CellMask™ Red, which is a plasma membrane stain, was co-incubated with ThT and viewed using confocal microscopy (see Supplementary Marerial).

### Fluorescence microscope

To be able assess the isolated “crescent pellet”, the PPP samples were centrifuged at 16,000 x g for 10 minutes at room temperature and then another 10 minutes at 0°C. The samples were stained with MPO and ThT (see Supplementary Material).

### Polarized microscope

Congo Red is commonly used in histochemical studies to detect amyloid and using a polarized microscope the Congo Red birefringence for amyloid turns apple green [51–53]. Samples were stained with Congo Red and ThT (see Supplementary Material).

### Scanning Electron Microscopy (SEM)

Whole-blood samples were processed for SEM on the day of collection, comprising fixation in 4% formaldehyde and 1% osmium tetroxide, graded ethanol dehydration, HMDS drying, and carbon coating (see Supplementary Material).

### Statistical analysis

For the TEG^®^ data, GraphPad Prism (version 10.5.0) was used for statistical analysis. Distribution normality was determined with the Sharpiro-Wilk test, after which data were either subject to Mann-Whitney or Welch’s t-tests. Statistical significance was determined at p < 0.05. Significance levels are indicated by asterisks (* = p < 0.05, ** = p < 0.01, *** = p < 0.001, and **** = p < 0.0001). Parametric data are presented as mean ± standard deviation (SD) and nonparametric data represented as median [IQR].

Two complementary and independent aspects of the flow cytometry data were analysed. *Abundance* (how many objects were detected by each experiment) was quantified as event counts, comprising both the total count per sample and the count within each of the six size bins. *Composition* (the shape of the size distribution, independent of abundance) was quantified as the proportion of a sample’s binned objects falling within each size bin; that is, whether the detected objects tended to be larger or smaller, irrespective of how many were present in total.

Control and Long COVID groups were compared using the two-sided Mann-Whitney U test (Wilcoxon rank-sum). A rank-based test was used because the count data were heavily right-skewed and contained both outliers and exact zeros, conditions under which the Mann-Whitney test remains robust and is unaffected by a pseudo-log_₁₀_ transform applied for display. The null hypothesis was that the two groups shared the same distribution, with no shift in either direction, rather than a directional Long COVID > Control hypothesis; the direction of any difference was inferred from the group medians. For each comparison, a Welch t-test was additionally computed as a parametric cross-check.

Because absolute counts appeared more variable in Long COVID samples, a supplementary analysis assessed whether this represented a genuine increase in dispersion or merely followed from the higher counts, since variance commonly scales with the mean. Rather than comparing the groups at the median alone, their full distributions were contrasted using a quantile-by-quantile comparison that shows how the difference between the groups changes across the distribution instead of assuming it is constant. Group quantiles were estimated with the Harrell-Davis estimator and their differences (Long COVID minus Control) taken at each decile, with 95% confidence intervals from a percentile bootstrap (2,000 resamples). The same quantiles were additionally expressed as ratios (Long COVID divided by Control) to separate an additive difference from a multiplicative one. Dispersion was then tested directly with the Fligner-Killeen test of homogeneity of variances, applied to both the raw counts and the log-transformed counts and summarised with scale-free measures of spread (the coefficient of variation and the log-scale standard deviation); a variance difference present on the raw scale but absent after log-transformation would indicate that the greater absolute spread in Long COVID followed from the higher counts (mean–variance coupling) rather than reflecting an independent increase in heterogeneity.

For the composition analysis, the six size-bin counts of each sample were converted to within-sample proportions, each bin being divided by the sum of the six bins. This adjustment removes sample-to-sample differences in the total amount of material and isolates the shape of the size distribution.

To summarise each sample’s size distribution in a single value, a size-shift index was computed as the count-weighted mean size-bin rank. The six size bins were assigned ordinal ranks from 1 (smallest) to 6 (largest), and the index was calculated as

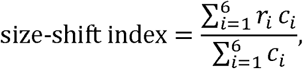

where *r*_i_ is the rank of bin *i* and c*_i_* its count. The index represents the average size class occupied by a sample’s objects: values near 1 indicate concentration in the smallest bins and values near 6 concentration in the largest, so a higher index in one group denotes a size distribution shifted toward larger objects. Because the weights are within-sample counts, the index depends only on the relative size distribution and not on the total number of objects detected, placing it within the composition rather than the abundance analysis. It complements the per-bin proportion comparisons by capturing the overall direction of any size shift in a single value. Ordinal bin ranks were used in preference to physical bin midpoints to avoid dependence on the arbitrary choice of a representative size within each bin.

The size composition was examined further using compositional data analysis, in which each sample’s six-bin proportion vector was treated as a point on the simplex. Exact zeros, which were infrequent (approximately 4% of bin entries), were addressed by adding a pseudo count of 0.5 to each bin count before transformation. Each proportion vector was then transformed using the centred log-ratio (CLR). For a composition x=(x_1,_ ,x_6_) of the six bin proportions,

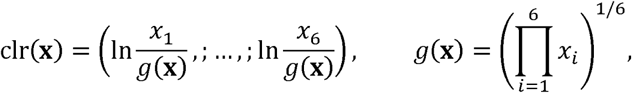

where *g*(x) is the geometric mean of the six proportions. Each bin is thereby expressed as the log of its proportion relative to the sample’s geometric mean, which removes the constant-sum constraint of the raw proportions and places them in an unconstrained real space suitable for standard distance and variance methods. The Aitchison distance between two samples was then taken as the Euclidean distance between their CLR vectors. For each experiment, differences in mean composition between Control and Long COVID were tested by permutational multivariate analysis of variance (PERMANOVA^1^) on the Aitchison distance matrix, using 9,999 permutations. Because a significant PERMANOVA can arise from a difference in group dispersion rather than in location, the homogeneity of multivariate dispersions was tested separately for each experiment through a permutation test of multivariate homogeneity of group dispersions (PERMDISP^2^) with 9,999 permutations, which compares the mean distance from samples to their group centroid and thereby tests directly whether compositional heterogeneity differed between groups. Compositional structure was visualised by principal coordinates analysis of the Aitchison distances, with 68% normal-probability ellipses drawn per group. As a targeted contrast of large against small objects, a log-ratio balance was also computed for each sample as the natural logarithm of the ratio of the geometric mean proportion of the three larger bins to that of the three smaller bins, with higher values indicating relatively greater representation of larger objects; balances were compared between groups with the Mann-Whitney U test and Benjamini-Hochberg correction across the five experiments [54].

Two further analyses quantified the magnitude and precision of any compositional difference. First, a directional effect size, Cliff’s delta [55], was computed for each size bin as the difference between the probability that a Long COVID value exceeds a Control value and the probability of the reverse; values range from −1 to +1, the sign indicating which group had the higher proportion in that bin, and 95% confidence intervals were obtained by bootstrap resampling (2,000 resamples). Second, an equivalence test was applied to the size-shift index to distinguish an absence of evidence for a difference from evidence for the absence of a meaningful difference. The between-group shift in the index was estimated using the Hodges-Lehmann estimator (the median of all pairwise Control minus Long COVID differences) with a 95% confidence interval, and equivalence was concluded when this interval fell entirely within a pre-specified margin. The margin was defined as a smallest effect size of interest on the ordinal rank scale, on which one unit corresponds to one full size class; the primary margin was set at ±0.5, that is half a size class, as the largest difference considered practically negligible. As no field-standard margin exists for this measure, the equivalence conclusion was additionally evaluated across a range of candidate margins (±0.3, ±0.5, ±0.75 and ±1.0 bins) to establish its robustness, with experiments equivalent across the whole range regarded as robustly equivalent and those whose conclusion changed with the margin flagged as margin-sensitive.

Multiple comparisons were controlled using the Benjamini-Hochberg false discovery rate, reported as q. Corrections were applied within families defined as the six size bins of each experiment; total (“All”) counts were treated as a single primary endpoint per experiment and reported unadjusted, and the size-shift index was corrected across the five experiments. PERMANOVA and PERMDISP were performed once per experiment and reported without adjustment across experiments. Significance was denoted ns (q ≥ 0.05), * q < 0.05, ** q < 0.01, and *** q < 0.001.

All analyses and figures were produced in R (v4.5.2). The Mann–Whitney U, Welch t-, and Benjamini–Hochberg procedures were computed with the base *stats* package. Additional packages were used for specific analyses: *vegan* (v2.7-2) for PERMANOVA and PERMDISP; *Hmisc* (v5.2.4) for Harrell–Davis quantile estimation; and *rstatix* (v0.7.3) with *ggpubr* (v0.6.3) for in-figure test annotation. Figures were drawn with *ggplot2* (v4.0.1), *patchwork* (v1.3.2), and *scales* (v1.4.0); data handling used *dplyr* (v1.1.4) and *tidyr* (v1.3.1).

## Results

### Thromboelastography (TEG^®^)

In order to assess the coagulation profile of the control and Long COVID cohort, both whole-blood and PPP samples were subject to TEG^®^ analyses. Significant differences were observed in five of the seven parameters assessed across both whole-blood and PPP samples, all of which pertain to clotting kinetics (Table 1). In the whole-blood analysis, Long COVID samples exhibited significantly shortened reaction time (R) (****) and clot formation time (K) (***), alongside an increased α-angle (**). Maximum rate of thrombus generation (MRTG) was also significantly elevated, while time to maximum rate of thrombus generation (TMRTG) was significantly reduced. These findings indicate accelerated clot initiation and propagation, and hence emphasize hypercoagulability in the Long COVID cohort. In contrast, maximum amplitude (MA) and total thrombus generation (TTG), which refer to the clot strength and amount formed, respectively, did not differ between groups, highlighting increased clotting kinetics despite normal clot strength and overall mass.

**Table 1:** TEG^®^ results of both whole-blood and PPP samples. Data are represented as mean ± SD, or median [IQR].

| TEG® of Whole Blood |  |  |  |
| --- | --- | --- | --- |
| Parameter | Control | Long COVID | P-value |
| R | 12.9 [11.2-15.4] | 9.2 [7.9-10.78] | **<0.0001** |
| K | 3.8 [3.1-4.4] | 2.5 [2.2-3.1] | **0.0002** |
| α-Angle | 39.1 [28.7-44.8] | 53.9 [41.4-56.9] | **0.0026** |
| MA | 59.5 ± 4.6 | 60.3 ± 5.4 | 0.61 |
| MRTG | 6.5 ± 2.0 | 8.4 ± 2.2 | **0.0068** |
| TMRTG | 16.2 [14.1-19.3] | 11.1 [9.4-13.2] | **<0.0001** |
| TTG | 715.7 ± 57.6 | 723.1 ± 64.5 | 0.71 |
| TEG® of PPP |  |  |  |
| R | 17.3 [13.2-26.0] | 9.4 [8.2-11.0] | **<0.0001** |
| K | 4.1 [3.1-5.2] | 2.6 [1.9-3.4] | **0.0073** |
| α-Angle | 20 [14.1-40.2] | 54.2 [38.6-62.1] | **0.0001** |
| MA | 29.3 [24.9-36.3] | 25.5 [23.4-36.6] | 0.29 |
| MRTG | 5.9 [4.3-7.2] | 9.8 [7.7-11.5] | **0.0002** |
| TMRTG | 19.8 [14.8-27.9] | 10.6 [9.2-12.3] | **<0.0001** |
| TTG | 350.6 [297.5-435.6] | 305.2 [280.6-438.3] | 0.29 |

The hypercoagulable phenotype of whole-blood was conserved in PPP as indicated by significant differences in all five kinetics-related parameters, albeit with different degrees of significance for K (**), α-angle (***), and MRTG (***). Again, MA and TTG were comparable between control and Long COVID groups. These results, specifically the persistence of increased clotting kinetics in PPP samples, are suggestive of hypercoagulability stemming, in a large part, from dysregulated coagulation factors and other plasma constituents.

### Imaging Flow Cytometry

FMCs are heterogeneous, spanning a continuum from cell-derived material to amyloid-bearing aggregates. To quantify this pool, imaging flow cytometry was used to measure three markers in PPP, comparing the control and Long COVID cohorts: ThT, reporting the amyloid-bearing fraction; Hoechst, reporting DNA-bearing cell-derived material; and CD62P-PE (P-selectin), reporting activated platelets (Figure 1). Long COVID PPP contained significantly more ThT-positive events than controls across all three sample preparations – undiluted PPP (*), 10x-diluted PPP (**), and the resuspended pellet (*) – indicating a greater burden of amyloid-bearing FMCs that persisted regardless of sample processing. Activated-platelet material (CD62P-PE-positive) was likewise elevated in Long COVID (**). By contrast, the broader pool of DNA-bearing, cell-derived material (Hoechst-positive) did not differ between the groups.

**Figure 1:**
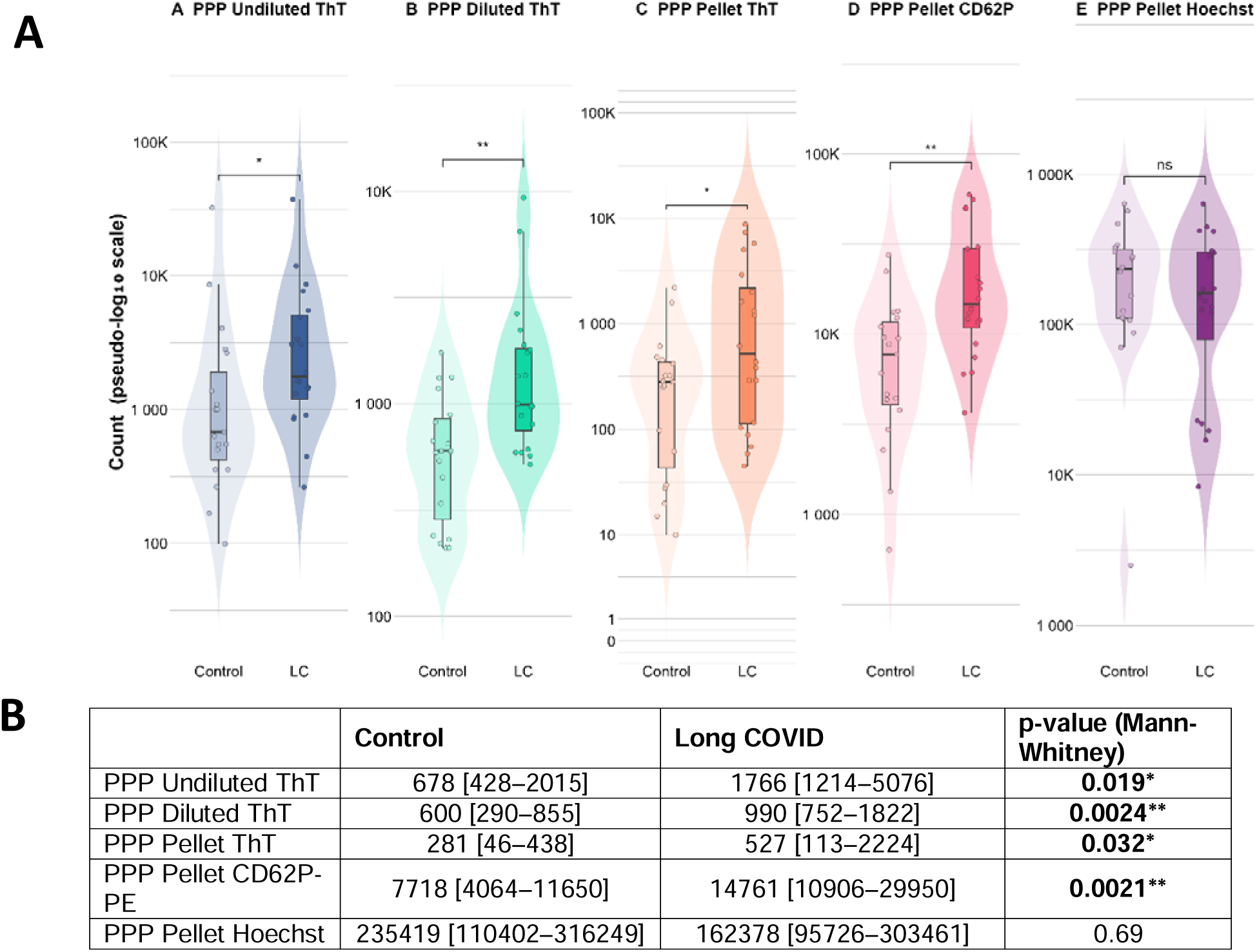
Flow cytometry results of undiluted PPP, diluted PPP, and resuspended PPP pellets stained with either ThT, CD62P-PE antibody-, or Hoechst. A) Box and whisker plots representing each control vs disease group pair. B) Table of count data with p-values for each comparison. Both box and whisker plots have had y-axis data transformed by log10. Data is represented as median [IQR]. LC = Long COVID. Brackets: Mann-Whitney U, BH-FDR q across the 5 methods (* q<0.05, ** q<0.01, *** q<0.001).

Between the three probes exposed to resuspended PPP pellets, nuclear material (Hoechst-positive signal) was shown to be the most abundant event type measured with a median of 235419 [110402–316249] and 162378 [95726–303461] for the control and Long COVID groups, respectively. Platelet material was the second most abundant event type with a median of 7718 [4064–11650] for the controls and 14761 [10906–29950] for the Long COVID group (**). Lastly, FMCs were the least abundant event type measured out of the three probes, with a median of 281 [46–438] and 527 [113–2224] for the control and Long COVID groups, respectively (*).

These elevated counts were also more variable in Long COVID, but this reflected mean-variance coupling rather than an independent increase in heterogeneity: the greater absolute spread scaled with the higher counts, and once the counts were log-transformed the between-group difference in variance was non-significant for every experiment (Fligner–Killeen), with the log-scale standard deviation near unity. On this basis the elevation is best characterised as an approximately multiplicative up-shift rather than a widening of the distribution, a pattern most clearly seen in the undiluted PPP and CD62P-stained samples and broadly consistent across the others (quantile-difference and quantile-ratio analyses; See supplementary Document).

Between sample types within either the control or Long COVID group, the median count of FMCs was highest in undiluted PPP and lowest in the resuspended pellet fractions (Figure 1). Although there was a loss in FMC detection upon either dilution or pellet obtainment, statistically significant group differences still remained when comparing the control and Long COVID groups. However, correlation analyses revealed no positive concordance between paired sample types within a given group; the only significant association was a negative correlation between the diluted and pellet ThT counts in the Long COVID group (Spearman ρ = −0.62, p = 0.003; Supplementary Material Table S5), indicating variation in FMC quantification between unprocessed and processed PPP samples. To add to this ambiguity is the fact that a large proportion of FMCs are still contained within the supernatant following centrifugation; this suggests that FMCs vary in density and hence obtaining a PPP pellet via the current centrifugation protocol will not isolate the entire population of ThT-positive events within plasma. ThT-positive events retained in the supernatant did not differ between the control and Long COVID groups (420 [305–787] and 410 [330–690], respectively), even though the undiluted, diluted and pelleted fractions all did.

Representative ThT images of undiluted PPP, diluted PPP, and the resuspended pellet are shown in Figure 2. Representative images of CD62P-PE- and Hoechst-positive events are shown in Figure 3A and B, respectively; morphological features of the ThT-, CD62P-PE-, and Hoechst-positive populations are considered in the Discussion

**Figure 2:**
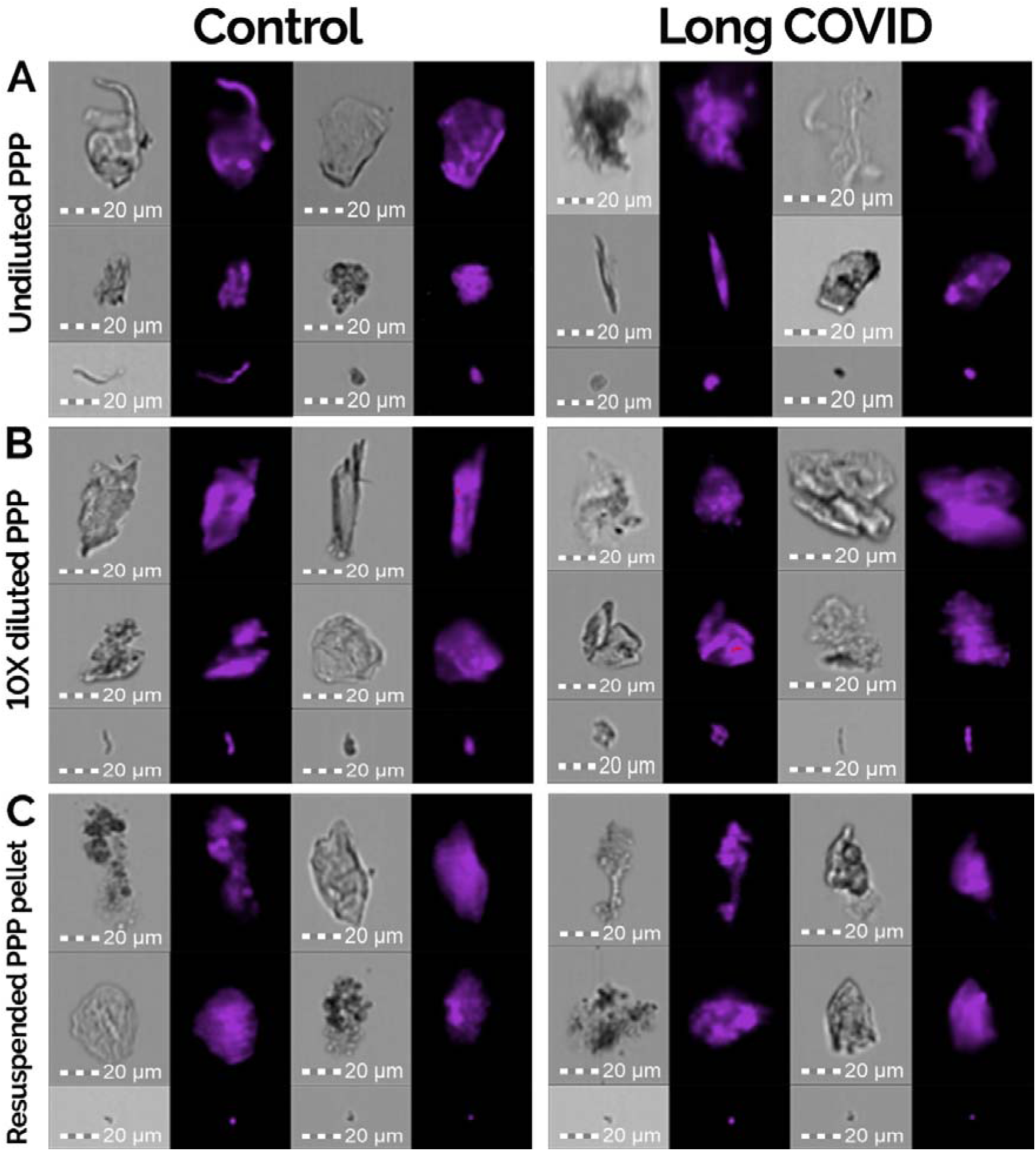
Images obtained from the flow cytometer of ThT-stained objects (purple); undiluted PPP (A), 10X diluted PPP (B), and the resuspended PPP pellet (C). The two image channels displayed are the brightfield (left) and fluorescence channel (right). Images were obtained using a 20X objective.

**Figure 3:**
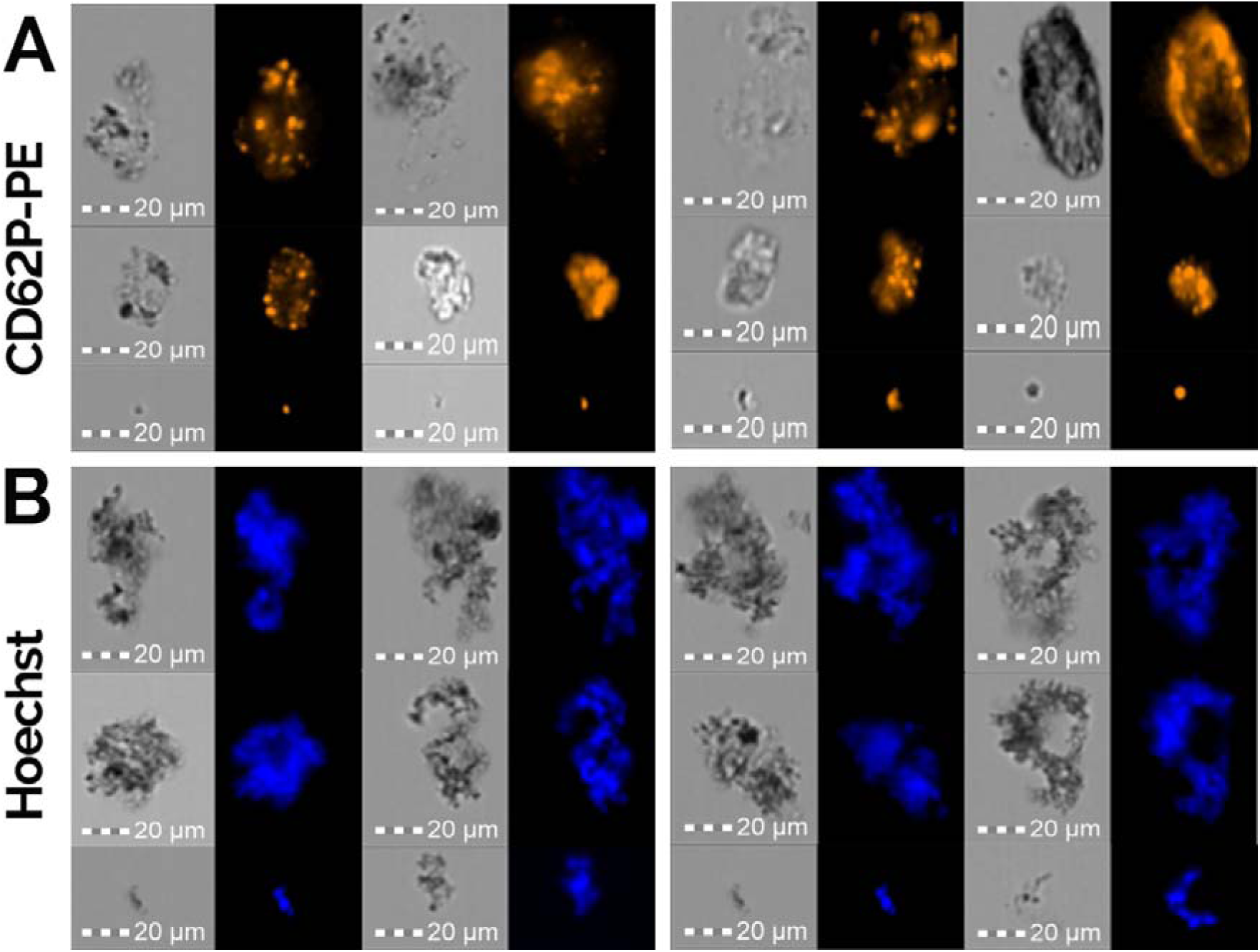
Images of CD62P-PE-positive events (A) and Hoechst-positive events (B). The two image channels displayed are the brightfield (left) and fluorescence channel (right). Images were obtained at 20X magnification.

To test whether Long COVID alters the *size* of these events rather than simply their number, each event population was expressed as its distribution across the six area bins and compared between groups (Figure 4A; full statistics in Supplementary document; Figures S5 to S9 and Tables S2 to S11). The size composition was largely preserved: the count-weighted mean size (size-shift index) did not differ between groups for any experiment after correction, and formal equivalence testing confirmed that the size distributions of diluted PPP ThT, pellet ThT, and pellet Hoechst events were statistically equivalent between control and Long COVID (Figure 4B; the 95% confidence interval for the shift fell within half a size bin). For these experiments, the increased event numbers in Long COVID are not accompanied by any change in size profile.

**Figure 4:**
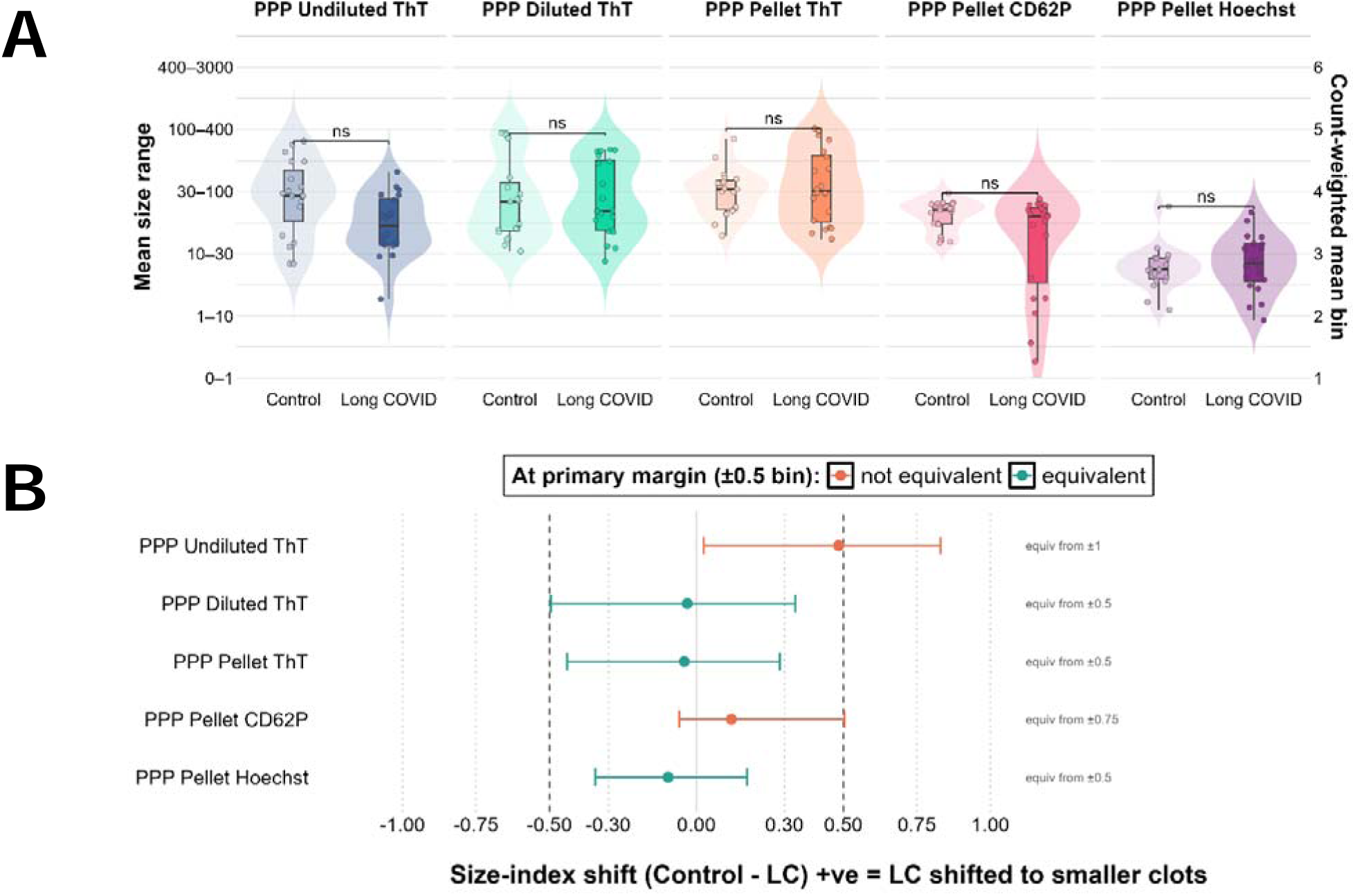
Size-shift index and its equivalence. (A) Size-shift index: the count-weighted mean size bin per sample (1 = smallest, 0–1 µm²; 6 = largest, 400–3000 µm²), comparing control and Long COVID groups for each experiment; higher values indicate a distribution shifted toward larger events. Mann–Whitney U with Benjamini–Hochberg correction across the five experiments; no comparison remained significant after correction. Data are median [IQR]. (B) Equivalence of the between-group size-index shift: the Hodges–Lehmann shift (control − Long COVID) with 95% confidence interval for each experiment, evaluated against candidate equivalence margins (vertical lines at ±0.3, ±0.5, ±0.75 and ±1.0 bins). An experiment is equivalent when its interval falls entirely within a margin (teal = equivalent at the primary ±0.5-bin margin, coral = not): Diluted ThT, Pellet ThT and Hoechst are equivalent from ±0.5 upward, CD62P only from ±0.75, and Undiluted ThT not until ±1.0.

Two experiments were exceptions (Figure 5). In undiluted-PPP ThT, the overall composition differed modestly (PERMANOVA p = 0.039), reflecting a suggestive shift toward *smaller* events in Long COVID; because this shift fell to trend level after correction and was accompanied by greater variability in the control group, we interpret it cautiously. In pellet samples stained with CD62P-PE, the average composition did not differ, but Long COVID samples were markedly *more heterogeneous* in their size distribution (PERMDISP p = 0.009), a compositional counterpart to the elevated activated-platelet counts. Overall, the Long COVID phenotype is therefore predominantly one of *more events of comparable size* rather than larger ones, with a suggestive skew toward smaller ThT events in undiluted plasma and greater heterogeneity of platelet-associated material.

**Figure 5:**
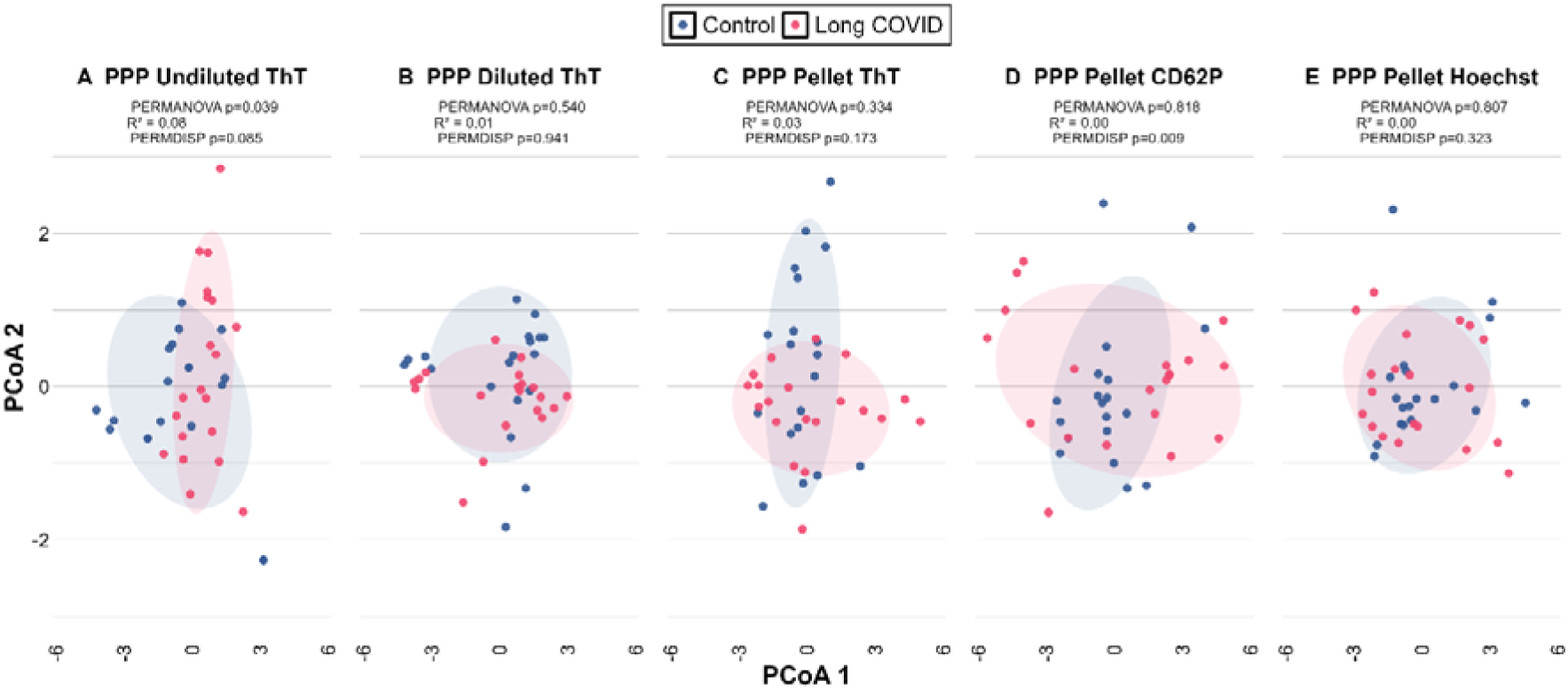
Compositional ordination. Principal coordinates analysis of the six-bin size composition (Aitchison distances), per experiment (A–E), comparing control and Long COVID groups. Each point is one sample; shaded ellipses are 68% normal-probability contours. PERMANOVA tests whether mean composition differs between groups and PERMDISP whether compositional dispersion (heterogeneity) differs (9,999 permutations each; statistics inset per panel). Undiluted ThT (A) shows a modest between-group compositional difference (PERMANOVA p = 0.039), and Pellet CD62P-PE (D) shows greater dispersion in Long COVID (PERMDISP p = 0.009); the other experiments overlap.

Because the three stains were applied to the same resuspended pellet (an identical participant set), the *size composition* of the amyloid-bearing (ThT), cell-derived (Hoechst), and platelet-associated (CD62P-PE) fractions can be compared directly within the FMC pool (Figure 6 and Table 2); the comparison is of within-sample proportions and is therefore independent of the stains’ differing absolute sensitivities. Each stain occupied a distinct size signature (Friedman p < 0.05 in every area bin, both cohorts). Cell-derived (Hoechst) material was concentrated in the smallest bins, exceeding both the amyloid-bearing and platelet-associated fractions at 1-10 µm² (***) and remaining the largest fraction at 0-1 µm². The platelet-associated (CD62P-PE) fraction peaked at platelet size (30-100 µm²), where it exceeded both the amyloid-bearing and cell-derived fractions (***). The amyloid-bearing (ThT) fraction, in turn, dominated the largest bins (100-400 and 400-3000 µm²), exceeding the platelet-associated fraction throughout (***) and the cell-derived fraction at 100–400 µm² (***); the 400-3000 µm² difference reached significance in control (**) but not Long COVID groups).

**Figure 6:**
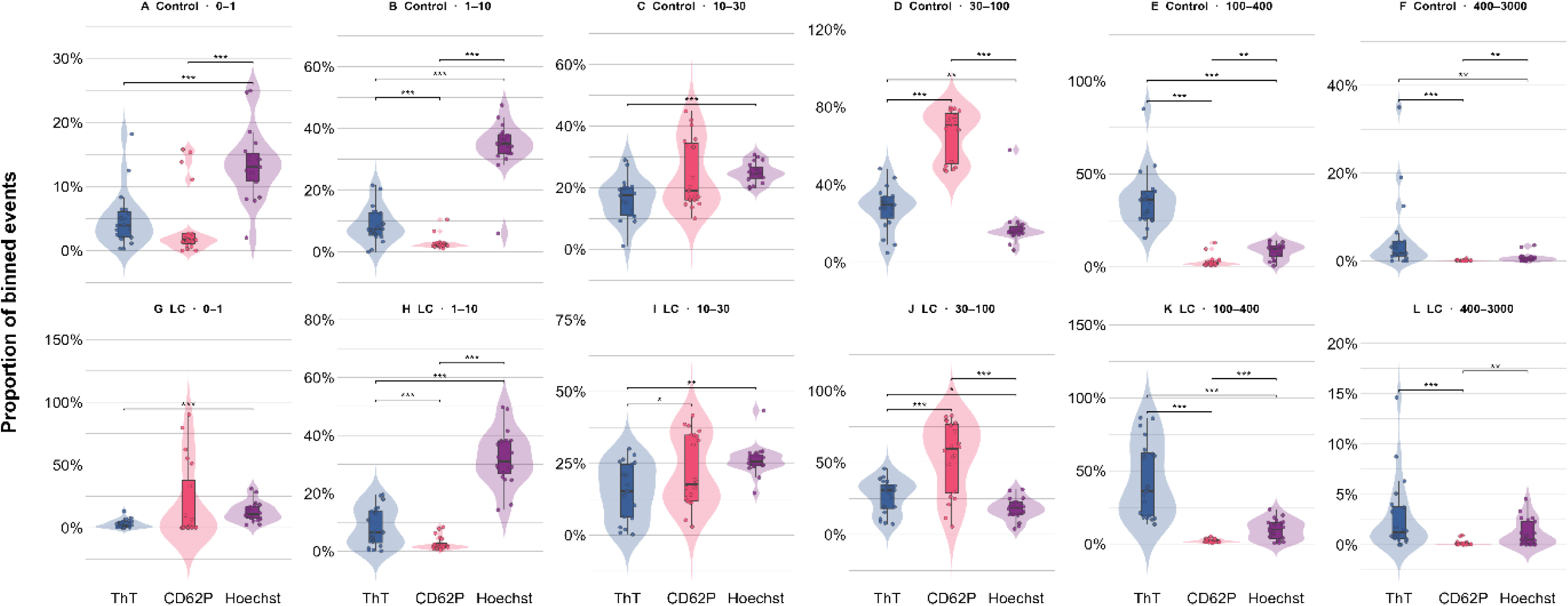
Distribution of ThT-, CD62P-, and Hoechst-positive events within each size range/area bin, shown for control (A–F) and Long COVID (G–L) groups. For each participant and size range, counts are expressed as a proportion of that sample’s binned total (violin = distribution, box = median/IQR, points = participants). Statistics: Friedman test per group x bin, with pairwise paired Wilcoxon signed-rank post-hoc (Benjamini–Hochberg-adjusted); brackets show significant comparisons (* q<0.05, ** q<0.01, *** q<0.001). (Additionally, see Table S5).

This ordering (cell-derived material smallest, platelet-associated material at platelet size, and amyloid-bearing FMCs largest) confirms that ThT reports a size-distinct amyloid-bearing subpopulation within the FMC pool, rather than merely restaining platelets or cell-derived particles. The sparse CD62P-PE-positive events in the two largest bins (where cell-derived material also exceeded them, **) most plausibly represent platelet aggregates or platelets complexed with leukocytes or other FMCs (Figure 3A).

### Confocal-, Fluorescence- and Polarized Microscopy

To assess the association of FMCs with cellular material, PPP samples were stained with ThT and Cellmask™ Red and then viewed using confocal microscopy (Figure 7A & B). Samples demonstrate considerable heterogeneity in FMCs: some FMCs display only ThT signal (Figure 7B) while others display both ThT and Cellmask™ Red signal (Figure 7A). This indicates that FMCs can exist with and without association to cellular membrane material. This cellular material can include platelet-, leukocyte-, erythrocyte-, and/or vesicle-derived structures. To further evaluate this, we added Congo Red in combination with either ThT or PAC-1 (Figure 7E-H). For both these combinations, the samples were positive for amyloidogenic and platelet debris that was associated with the Congo Red signal.

**Figure 7:**
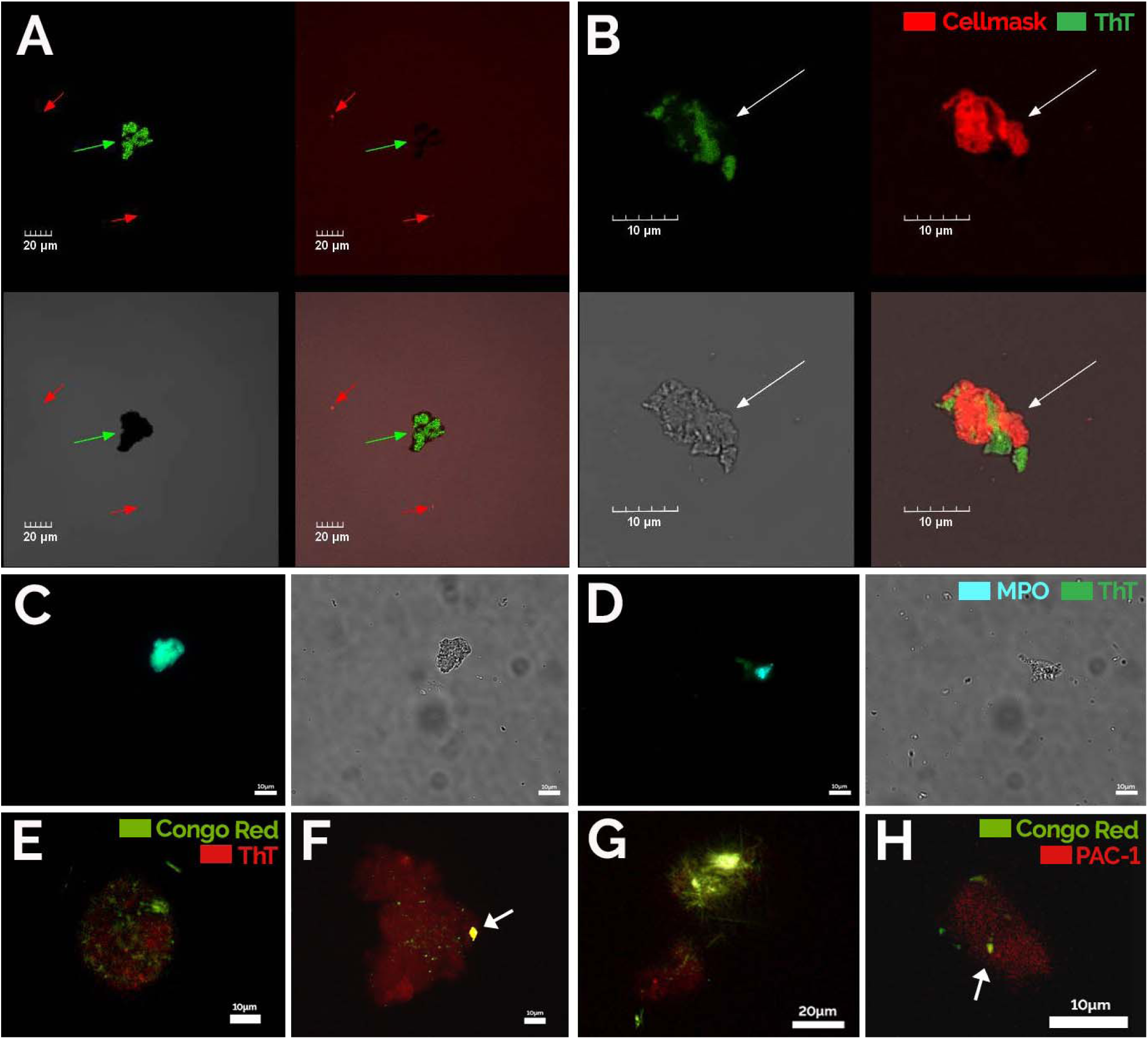
Confocal-, fluorescence- and polarized microscopy micrographs of PPP samples from the Long COVID cohort. **Figures A and B** are confocal micrographs of samples exposed to ThT (green fluorescence) and Cellmask™ Red (red fluorescence). The third micrograph in each grid block is representative of the transmitted channel and the last micrograph represent the overlay. **Figures C and D** represent fluorescence micrographs of the “crescent-shaped” pellet part exposed to MPO (blue fluorescence) and ThT (green fluorescence). **Figures E-H** represent FMCs exposed to Congo Red (apple green) (arrows) and (E-G) ThT (red fluorescence) (For F, arrow shows ThT and Congo Red co-staining as yellow); (H) Congo Red (apple green) and PAC-1 (red fluorescence). Imaged using both polarized and fluorescence microscopy. Scale bars: B, C, D, E, F, H: 10 µm; A, G: 20 µm.

During centrifugation of PPP at 16,000 x g, we noted a second insoluble fraction towards the top of the tube opposite the basal pellet (on the side of the tube closest to the centre of the centrifuge), consistent with a lipid fraction. This crescent-shaped deposit is referred to as the “crescent pellet”. This “crescent pellet” stained positive for, MPO and ThT (Figure 7C-D), indicating the presence of FMCs positive for neutrophil, and amyloid material. We report this observation simply to note that PPP appears to contain at least one further distinct insoluble fraction beyond the basal pellet after intensive centrifugation. The nature of this second insoluble PPP fraction in the context of FMCs, cell debris, and lipid content remains to be investigated.

### Scanning Electron Microscopy

High-resolution analysis of whole blood smears was performed using scanning electron microscopy (Figure 8). Control samples exhibit predominantly discoid erythrocytes with smooth membranes, a low abundance of activated platelets, and few regions of platelet clumping and/or debris. In contrast, whole-blood smears from the Long COVID cohort demonstrated marked platelet hyperactivation, characterized by extensive pseudopodia formation, platelet spreading, and frequent platelet aggregation. Furthermore, erythrocytes in the Long COVID cohort showed pronounced agglutination and close association with hyperactivated platelets, forming aggregates that were not frequently observed in controls. Few eryptotic RBCs were detected in either cohort, with no obvious qualitative differences between groups. These ultrastructural findings are in accord with the hypercoagulable profile (Table 1) and increase in platelet-derived debris (Figure 1) in the Long COVID group, together emphasizing sustained platelet activation, dysregulation, and a prothrombotic phenotype.

**Figure 8:**
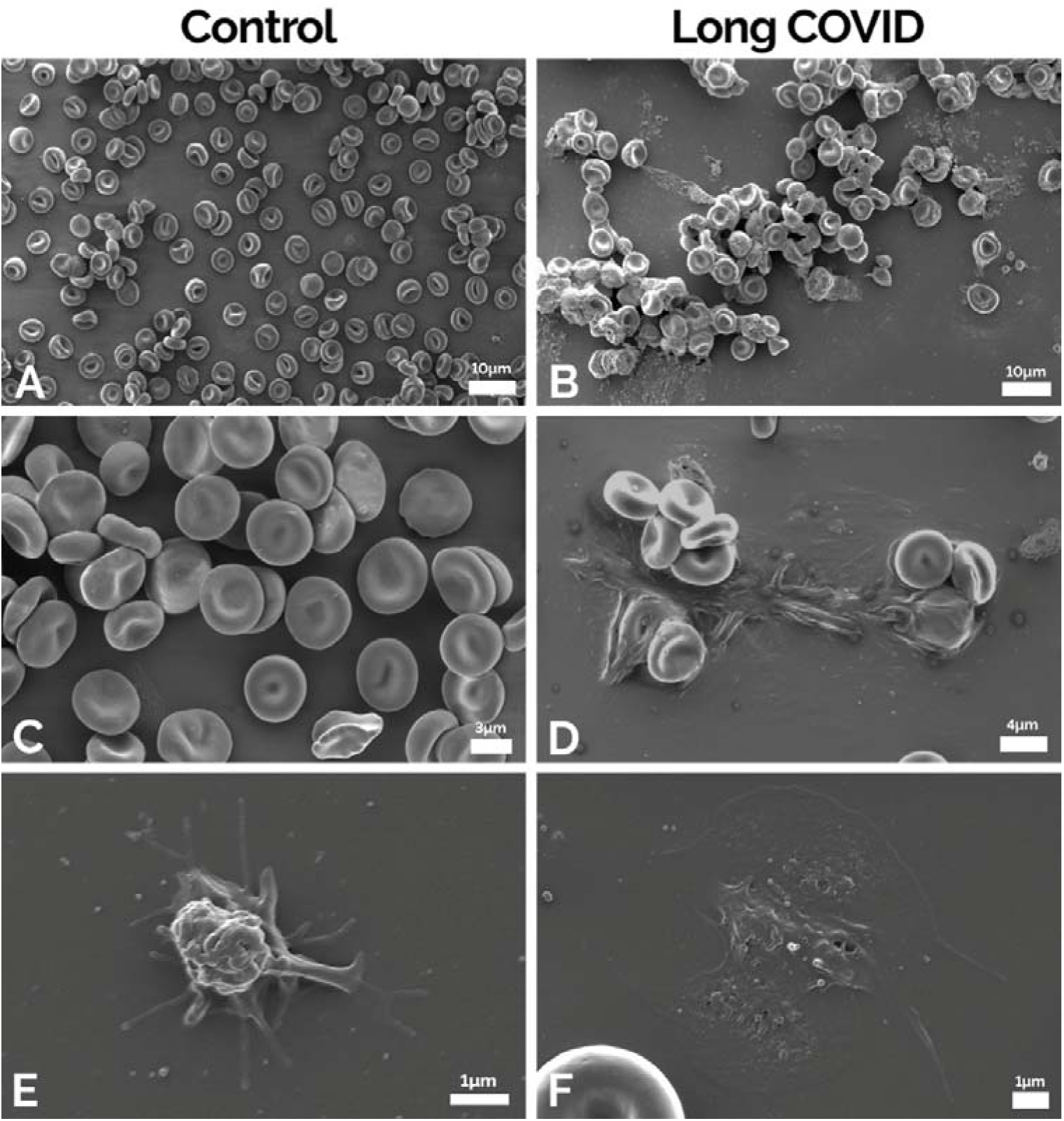
Scanning electron microscopy micrographs of control (A, C, E) and Long COVID (B, D, F) whole-blood samples. Control red blood cells and platelets are normally shaped with the control platelets only showing signs of contact activation (E). The Long COVID red blood cells show spontaneous clumping and activated platelets. Images were vied with the Zeiss MERLIN^TM^ field emission scanning microscope. All micrographs within a column represent different participants within a cohort. Arrows: Scale bars: A, B: 10 µm; C: 3 µm; D: 4 µm; E, F: 1 µm.

## Discussion

Hypercoagulation, platelet dysfunction, endothelial damage, and immune dysfunction, as well as increased levels of FMCs, are features of Long COVID pathology [9–11, 13, 14, 31, 56–59]. Interestingly, sustained prothrombotic changes have also been noted in a murine post-acute model [60], further supporting the persistence of prothrombotic sequelae after acute infection has been resolved. In our 2026 paper [24], we discussed the various FMC phenotypes, and proposed that prothrombotic cellular debris represent one of the seeding areas for FMCs. It was previously noted that plasma contains cell-derived debris that is itself proinflammatory [38].

While purified-fibrinogen systems where various inflammatory molecules are added, only studies a single FMC phenotype [27, 28, 36], (the fibrinogen and inflammatory molecule phenotype), the events observed in samples of whole blood or plasma are more heterogeneous. This heterogeneity is supported by data from proteomic analyses that indicate that FMCs contain fibrin(ogen) subunits, while being enriched in and entrapping a wide range of amyloidogenic and low-abundance plasma and cellular proteins [24, 31, 32, 34], as well as NETs [21]. Further elucidation of FMC biology is required, alongside standardization of their detection, classification, and quantification, to enable reproducibility across studies and in characterizing their mechanistic contributions to pathophysiology.

In this manuscript, we therefore assessed coagulation status, quantified FMCs across undiluted PPP, diluted PPP, and resuspended PPP pellets, and measured platelet- and nuclear-derived material within resuspended pellets in both control and Long COVID samples.

### Persistent Hypercoagulability in Long COVID

TEG^®^ was utilized to assess the state of the coagulation system in both whole-blood and PPP samples (Table 1). The results demonstrate extensive hypercoagulability in the Long COVID cohort, where significant differences were observed in all five parameters related to clotting kinetics. These differences persisted in PPP samples, which are devoid of intact platelets, as well as leukocytes and erythrocytes, suggesting that intrinsic plasma constituents significantly contribute to the procoagulant phenotype observed. These can include differences in fibrinogen levels, thrombin generation, inhibitors, fibrinolytic molecules, proinflammatory molecules, and other factors. PPP from Long COVID patients has been shown to promote the activation of healthy platelets [57], demonstrating that plasma constituents play a large role in amplifying clotting processes in this condition. We have also reported that sera from Long COVID patients significantly inhibit fibrinolysis in a purified fibrinogen model [17], further supporting the influence of plasma- and serum-derived factors in bringing about dysfunction in the coagulation system. Amyloid forms of proteins are of course well known to be more resistant to proteolysis [61]. However, this does not exclude a substantial contribution from platelets and other cellular components to the whole-blood TEG^®^ results, especially since the increased presence of platelet-derived material in PPP from the Long COVID cohort (Figure 1) further suggests ongoing platelet activation (despite a lack of positive correlations between CD62P-PE-positive counts and TEG^®^ parameters).

These TEG^®^ results reflecting increased clotting kinetics are consistent with other studies showing increases in prothrombin time [11], thrombin generation [56, 62], platelet dysregulation [8, 13, 57], and elevations in clotting factors [14, 58, 63, 64], but in contrast to a case-control study where TEG^®^ values were similar between control and Long COVID cohorts [65]. This discrepancy may reflect underlying variability in the coagulation phenotype of Long COVID patients and highlights the need for further clarification of TEG^®^ findings across larger cohorts. Continued and more detailed investigation into the mechanistic basis and clinical relevance of clotting dysfunction in Long COVID is warranted.

### Elevations in FMCs and Platelet Debris in PPP Samples

The measurement of FMCs was carried out using imaging flow cytometry. Indeed, subjecting PPP samples to flow cytometry is not routine, as intact cell populations are largely absent, but the ability of flow cytometers to detect events in PPP (such as cell debris and FMCs), which range in size from microns to hundreds of microns in area, warrants exploration of its application in this regard [26]. In this paper, FMC quantification was carried out on three different sample types, namely undiluted PPP, 10X diluted PPP, and resuspended PPP pellets. For flow cytometry, naive PPP can easily lead to blockages and impact overall quantification; hence, diluted PPP and resuspended PPP pellet fractions were also tested and measured against naïve PPP.

Between control and Long COVID groups, significant differences were observed across all sample types, with the Long COVID groups exhibiting more FMCs than the control groups (Figure 1). These findings further emphasize that elevations in FMCs persist in Long COVID, which may contribute to ongoing thrombo-inflammatory and vascular dysfunction. This is in accord with other studies that have quantified FMCs in plasma [20, 22, 23], albeit without the use of flow cytometry.

In addition to measuring FMCs with ThT, PPP pellets were also stained with a CD62P-PE antibody and Hoechst to quantify insoluble (prothrombotic) aggregates and FMCs that contain platelet and leukocyte debris. Before interpreting the comparisons between these populations, one constraint must be stated, because it determines what can and cannot be concluded: ThT, the CD62P-PE antibody, and Hoechst were applied to separate aliquots (not co-stained). Consequently, this flow cytometry quantification can establish the existence, abundance, size, and morphology of each labelled population independently, but it cannot resolve, at the level of an individual particle, whether platelet- or nuclear-derived events also carry amyloid character, or conversely whether ThT-positive events contain platelet or nuclear material (rather, this can be inferred although not quantified from Figure 7 where microscopy was used to assess co-stained samples). We are therefore careful to make population-level statements only, and to avoid any single-event co-localisation claim from the flow cytometry data.

In the resuspended PPP pellet, the Long COVID group exhibited a significantly higher median (14761 [10906–29950]) of CD62P-PE-positive events than the control group (7718 [4064–11650]) (**) (Figure 1). This increase in platelet debris in Long COVID PPP supports the hypercoagulable TEG^®^ results (Table 1) and likely reflects an increase in platelet activity (and hence clotting propensity). Because whole-blood samples were subject to coagulation assessments, platelet inhibitors were not applied to samples, and hence *ex vivo* platelet activation may have contributed to the platelet-derived material detected in PPP, particularly when whole-blood samples were centrifuged to obtain PPP. Nevertheless, standardized blood collection and processing procedures were applied consistently across all samples, enabling comparison between the control and Long COVID groups under equivalent experimental conditions.

No statistically significant differences in Hoechst-positive signal were observed between the control and Long COVID resuspended PPP pellet samples. It must be noted that the lack of significant differences in Hoechst-positive counts does not exclude a potential association of leukocyte material with FMCs in this flow cytometry analysis [21]. Although Hoechst staining reveals the presence of nuclear material within PPP, it is not specific for NETs or any particular leukocyte subset, and can stain both live and dead cells. Hoechst signal may therefore reflect a combination of NET-associated DNA, extracellular DNA including that of platelet mitochondrial DNA [66], and nuclear debris with leukocyte material.

Within the resuspended pellet samples, FMCs were the least abundant event type, while nuclear material-associated events were the most abundant, followed by platelet material (Figure 1). These data emphasize that ThT does not randomly and indiscriminately bind cellular debris, but instead the ThT-positive fraction presents a unique concentration from the majority of cellular debris as measured by the CD62P-antibody and Hoechst.

### Probe-Specific Events Differ in Size Distribution and Morphology

ThT-, CD62P-, and Hoechst-positive material occupied distinct size domains (Figure 6 and Table 2), with significant differences in size distribution between probes. Events with platelet and nuclear material predominated in the smaller size bins, namely 30–100 µm² and 1–10 µm², respectively, consistent with platelet and leukocyte-debris sizes; whereas ThT-positive-only FMCs were proportionally greater in the larger bins (100-400 and 400-3000 µm²), exceeding those FMCs sizes with platelet signal only and overlapping with FMCs carrying both leukocyte and platelet-leukocyte signal. [67]. FMCs with ThT-positive events only also outnumbered Hoechst-positive events in these bins. These data indicate a size-based organisation in which platelet and nuclear material dominates smaller size ranges and ThT-positive FMCs predominate in the larger ones, supporting the notion that there is a population of FMCs that is structurally distinct entities not reducible to platelet or leukocyte debris. These findings also underscore the need for an appropriate probe: FMCs have been detected using platelet markers such as CD62P and GPIIb/IIIa [68], but because platelet debris far outnumbers ThT-positive only FMCs and FMCs without membrane material exist (Figure 5A), platelet markers alone will misidentify the ThT-positive population. ThT remains the defining probe FMCs.

**Table 2:** Distribution of ThT-, CD62P-, Hoechst-positive events within the resuspended pellet, per area bin and cohort. Values are the median [IQR] percentage of each sample’s binned events falling in that bin, for ThT, CD62P-PE and Hoechst. p-values are pairwise paired Wilcoxon signed-rank comparisons (Benjamini–Hochberg-adjusted) following a significant Friedman test; bold denotes q < 0.05.

| Area Bin | Probe | Control | Long COVID | Comparison | Control p-value | Long COVID p-value |
| --- | --- | --- | --- | --- | --- | --- |
| 0–1 $\mu\text{m}^2$ | ThT | 3.9 [2.1-6.1] | 1.7 [0.3-4.8] | ThT vs CD62P | 0.42 | 0.58 |
|  | CD62P | 1.6 [1.1-2.6] | 0.2 [0.1-37.7] | ThT vs Hoechst | **<0.001** | **<0.001** |
|  | Hoechst | 13.1 [10.9-15.2] | 10.8 [7.4-17.1] | CD62P vs Hoechst | **<0.001** | 1.0 |
| 1–10 $\mu\text{m}^2$ | ThT | 7.4 [5.4-12.7] | 6.6 [3.0-14.0] | ThT vs CD62P | **<0.001** | **<0.001** |
|  | CD62P | 2.1 [1.9-2.9] | 1.8 [1.2-2.8] | ThT vs Hoechst | **<0.001** | **<0.001** |
|  | Hoechst | 35.0 [31.9-38.0] | 31.0 [26.7-38.1] | CD62P vs Hoechst | **<0.001** | **<0.001** |
| 10–30 $\mu\text{m}^2$ | ThT | 17.5 [11.1-20.1] | 15.3 [6.3-24.7] | ThT vs CD62P | 0.054 | **0.018** |
|  | CD62P | 19.0 [16.0-34.4] | 17.8 [11.9-34.9] | ThT vs Hoechst | **<0.001** | **0.004** |
|  | Hoechst | 24.5 [23.0-26.8] | 25.7 [24.3-28.0] | CD62P vs Hoechst | 0.77 | 0.25 |
| 30–100 $\mu\text{m}^2$ | ThT | 29.8 [22.8-34.0] | 30.7 [18.1-34.8] | ThT vs CD62P | **<0.001** | **<0.001** |
|  | CD62P | 70.8 [50.8-76.7] | 59.7 [28.9-76.5] | ThT vs Hoechst | **0.008** | **0.030** |
|  | Hoechst | 16.1 [14.6-18.4] | 18.5 [14.0-22.5] | CD62P vs Hoechst | **<0.001** | **<0.001** |
| 100–400 $\mu\text{m}^2$ | ThT | 36.1 [25.8-40.7] | 36.2 [18.9-62.4] | ThT vs CD62P | **<0.001** | **<0.001** |
|  | CD62P | 2.0 [1.3-3.0] | 1.6 [1.3-3.0] | ThT vs Hoechst | **<0.001** | **<0.001** |
|  | Hoechst | 9.9 [5.8-11.5] | 9.9 [3.6-14.7] | CD62P vs Hoechst | **0.004** | **<0.001** |
| 400–3000 $\mu\text{m}^2$ | ThT | 1.8 [1.1-4.4] | 1.2 [0.6-3.8] | ThT vs CD62P | **<0.001** | **<0.001** |
|  | CD62P | 0.1 [0.1-0.2] | 0.1 [0.0-0.1] | ThT vs Hoechst | **0.005** | 0.058 |
|  | Hoechst | 0.6 [0.1-0.9] | 0.5 [0.0-2.3] | CD62P vs Hoechst | **0.003** | **0.006** |

When looking at the images obtained from the flow cytometry experiments (Figure 2 and Figure 3), there is a morphological distinction between the events stained with Hoechst and those stained by either ThT and the CD62P-PE antibody. Hoechst-positive events displayed increased granularity in the brightfield images, characterised by irregular, textured structure and uneven signal density. These events frequently exhibited compact or clustered morphologies with uneven boundaries, consistent with the presence of nucleated or nuclear-associated material rather than membrane-bound, intact blood cells. As Hoechst labels DNA regardless of cell viability, it cannot distinguish free nuclear material from nuclear material still associated with cellular debris, so the relative proportions of each among the Hoechst-positive events remain unknown.

In contrast to the Hoechst-positive events, the morphologies of objects stained with both ThT and CD62P-PE events are more similar, and also unique from the Hoechst-stained events. Fluorescence associated with CD62P-PE was localised to the periphery of these events and frequently displayed a punctate or clustered distribution, consistent with surface localization rather than diffuse staining. The sizes of these punctate are also consistent with the size of platelets and platelet debris. These punctate were specifically noticeable on large events, particularly those similar in size to leukocytes. Associations between platelets and leukocytes, such as monocytes, have been demonstrated in Long COVID cohorts and have been shown to be elevated compared to controls [67]; the large CD62P-PE-positive events (>100µm^2^) with a punctate phenotype are proposed to represent these aggregates and debris thereof, but also FMCs. In contrast to ThT-positive events, which frequently exhibited amorphous or diffuse morphologies with poorly defined boundaries, CD62P-PE-positive events exhibited more prominent outlines.

### Association of FMCs with Cellular Material

Because only single-stained samples could be analysed via flow cytometry, microscopy was used to assess costained samples, enabling the identification of both cellular and amyloid material associated with FMCs (Figure 7). As shown in Figure 7A and B, FMCs were heterogeneous; some were ThT-positive only (lacking cellular material), while others were positive for both ThT and CellMask™ Red, indicating co-association of FMCs with cellular material. Extrapolating these findings to the flow cytometry data, it is important to note that while ThT, CD62P-PE, and Hoechst were analysed as single stains, there still exists the need to co-stain and determine the concentrations of PPP events positive for either or both amyloid and cellular material. Therefore, while the current flow cytometry data quantify the abundance of each marker independently, they do not permit conclusions regarding the co-localisation or physical association of ThT-positive FMCs with cellular material, which is apparent in Figure 7.

It was also demonstrated, using fluorescence microscopy (Figure 7C-D), that the “crescent pellet” comprised a heterogeneous mixture of biological material, including FMCs (ThT-positive) associated with cellular material (MPO-positive). The co-occurrence of these markers indicates that the “crescent pellet” contain amyloid proteins and cellular material, representing another complex cellular and proteinaceous fraction. This was also shown in the evaluation of PPP with Congo Red in combination with ThT or PAC-1 (platelet marker) (Figure 7E-H). The presence of Congo Red-positive material further corroborates the ThT signal, providing additional evidence that FMCs within plasma exhibit amyloid characteristics and also contains platelet debris. The presence of both amyloid (including fibrin(ogen)) and cellular material in FMCs likely contribute to the fibrinolytic resistance exhibited by these aggregates [27, 30, 32, 49], and hence can inform prospective experiments and therapies that aim to degrade these complexes, i.e. plasmin alone is thus likely insufficient to effectively reduce FMC burden *in vivo*.

Building on previous insights [24], damaged or dying cells and other cellular material in circulation that has not yet accumulated amyloid proteins are not to be considered inert. The exposure of PS on damaged cell membranes provides a highly procoagulant surface capable of promoting thrombin generation, fibrin deposition, and the recruitment of additional cellular and proteinaceous material [24]. PS externalization occurs during apoptosis, necrosis, platelet activation, and other forms of cellular stress, creating a scaffold that facilitates coagulation even before amyloid fibrin formation has occurred. As such, these ThT-negative aggregates may represent early intermediates in the evolution of FMCs, serving as induction sites that progressively accumulate fibrin(ogen), inflammatory proteins, and other blood components before ultimately adopting the β-sheet-rich amyloid conformation detectable by ThT or Congo Red. This proposed continuum suggests that pathological membrane remnants and cellular debris are not separate from FMC biology, but rather constitute precursor structures that may mature into the ThT-positive FMCs.

### Potential Contribution of Platelet-Derived Amyloid-β to ThT Signal in Blood

As noted in the introduction, platelets store pre-formed Aβ peptides, including Aβ_₁- ₄₂_, within α-granules, which are rapidly released upon activation by thrombin, LPS collagen, and arachidonic acid [41, 42]. As noted with fluorescence microscopy, platelet cellular debris fragments show both ThT and Congo Red signal, which might be due to the presence of Aβ. This platelet-derived Aβ has the potential to contribute to the amyloidogenicity of FMCs derived from platelet cell debris. Further investigation of the role of platelet-derived Aβ in FMC formation is therefore warranted.

### Conclusion and Future Directions

Overall, the differences in the concentrations of ThT-, CD62P-PE-, and Hoechst-positive events in both the control and Long COVID groups likely do not simply reflect differences in the abundance of distinct PPP aggregates, but rather represent different stages along a continuum of FMC formation. As ThT-positive FMCs were present at significantly lower concentrations than CD62P-PE- and Hoechst-positive events, these findings suggest that only a subset of cellular material progresses to form ThT-positive FMCs, indicating that the acquisition of an amyloid phenotype is likely a relatively late event in the process of FMC formation. If only a proportion of cellular material ultimately develops into FMCs, the nature of these precursor events and the mechanisms governing their transition into amyloid-positive FMCs warrant further investigation.

Ultimately, this study highlights the hypercoagulable profile of Long COVID patients and demonstrates comparative differences between the concentrations of FMCs and that of aggregates or FMCs containing platelet and leukocyte material. The Long COVID cohort exhibited significantly elevated levels of both ThT-positive FMCs across all sample preparations as well as CD62P-PE-positive (platelet) material.

In PPP, ThT-positive-only FMCs are numerically minor relative to and morphologically distinct from aggregates or FMCs that contain platelet- and leukocyte-derived material, specifically when single stains are used. The lack of indiscriminate ThT binding supports the specificity in detection of FMCs using ThT. Microscopy imaging of co-stained samples confirms that a subset of FMCs is membrane-free, establishing the existence of amyloid-rich, proteinaceous events. Importantly, because ThT and cell-specific probes were not co-applied in the flow cytometry experiments, this study makes no claim about the amyloid status of cellular debris in either direction; subsequent studies need to implement co-staining and methodological standardisation when using PPP in flow cytometry experiments.

### Limitations of Using PPP in Flow Cytometry

Applying flow cytometry to PPP is unconventional. The wide size range of events precludes narrow gating and complicates masking and fluorophore titration, increasing the risk of over-or under-estimating sizes (although the broad inclusive gates employed limit exclusion bias). Although undiluted PPP was assessed in this study, diluted or resuspended-pellet samples are preferable, especially for compatibility with flow cytometers. Standardised protocols for preparation, dilution, instrument settings, and gating are needed for reproducibility. With such standardisation, using PPP in flow cytometry experiments offers a novel route to study FMCs and cell debris in plasma samples.

Reassuringly, the broad inclusive gating did not appear to bias the group comparison: between-group size distributions were statistically equivalent for the diluted-PPP, pellet-ThT and pellet-Hoechst preparations, and the between-group variance was non-significant for every experiment after log-transformation (Fligner–Killeen), so the elevated Long COVID counts are unlikely to be an artefact of size-dependent gating.

#### Ethical clearance

Ethical clearance was granted by the Health Research Ethics Council (HREC) of Stellenbosch University, South Africa. This study (project ID: #34446) made use of anonymous samples collected under the Stellenbosch University Blood Laboratory Biorepository (project ID: #28804). All Biorepository samples were obtained with voluntary informed consent, including consent for storage and future use in approved studies. All research protocols were conducted in accordance with the Declaration of Helsinki, the South African Guidelines for Good Clinical Practice, and the Ethical Guidelines for Research issued by the South African Medical Research Council.

#### Author Contributions

MN: Conducted experimental analyses, wrote the paper; CMPG: Conducted experimental analyses, editing of manuscript; J-HP: Statistical analysis and editing of manuscript; CV: Imaging and editing of manuscript; BCF: Editing of manuscript; DBK: Editing of manuscript. EP: Editing of manuscript, funding, co-corresponding author; study leader. All authors have read and agreed to the published this version of the manuscript.

## Supporting information

All supplementary figures

## Data Availability

All data produced in the present study are available upon reasonable request to the authors

## Acknowledgements and Funding

M.N. thanks Kanro Foundation, E.P. thanks PolyBio Research Foundation, Balvi Foundation and Kanro Foundation for funding. The content and findings reported and illustrated are the sole deduction, view and responsibility of the researchers and do not reflect the official position and sentiments of the funders. The funders had no role in study design, data collection and analysis, decision to publish, or preparation of the manuscript.

## Declaration of generative AI and AI-assisted technologies in the manuscript preparation process

During the preparation of this work, the authors used Claude for language checks and review code. The authors reviewed and edited the output as needed and take full responsibility for the content of the published article.

## Footnotes

1 Using the adonis2 function in vegan.

2 Using the betadisper function in *vegan*.

