## Supplementary material for "Persistent Hypercoagulability and Further Characterization of Microclot Complexes in Long COVID": All supplementary figures

**Ethical clearance**

Ethical clearance was granted by the Health Research Ethics Council (HREC) of Stellenbosch University, South Africa. This study (project ID: #34446) made use of anonymous samples collected under the Stellenbosch University Blood Laboratory Biorepository (project ID: #28804). All Biorepository samples were obtained with voluntary informed consent, including consent for storage and future use in approved studies. All research protocols were conducted in accordance with the Declaration of Helsinki, the South African Guidelines for Good Clinical Practice, and the Ethical Guidelines for Research issued by the South African Medical Research Council.

**Sample cohorts and collection**

Healthy control individuals were included as such if they were without disease, did not smoke, were not using anticoagulant, antiplatelet, or anti-inflammatory medication, and had not reported a known infection and/or received a vaccination within a month of sample collection. All Long COVID patients were diagnosed by a physician at Stellenbosch Mediclinic. For patients to have been diagnosed with Long COVID, symptom onset must have occurred following acute COVID-19 infection and persisted for more than 3 months. Symptoms included in the diagnostic assessment were brain fog, chronic fatigue, post-exertional malaise, joint and muscle pain, gastrointestinal issues, and sleep disturbances. No patients were admitted to the hospital during acute SARS-CoV-2 infection. Of the 20 Long COVID participants, 17 reported having received at least one SARS-CoV-2 vaccination. Figure S1 and Table S1 contain the age distributions and other demographics of the sample cohorts. Blood samples were collected by a qualified medical professional in sodium citrate blood tubes (3.2%; BD Vacutainer^®^: 369714). A fraction of whole-blood obtained was assessed and the rest was centrifuged at 3000 x g for 15 minutes at room temperature. PPP was carefully collected, aliquoted into 1.5mL microcentrifuge tubes, and analysed or stored at -80ºC until further use.

**Table S1:** Demographics of the two cohorts, including comorbidities and symptoms of the Long COVID group. Out of the 20 Long COVID participants, half were afflicted with at least one other condition. Data were assessed with Mann Whitney tests and are presented as median [IQR].

| **Ages of the Cohorts** | | | |
| --- | --- | --- | --- |
|  | Control | LC | P Value |
| Age | 30 [20] | 40 [11] | 0.06 |
| Sex | 14 Females, 5 Males | 13 Females, 7 Males | N/A |
| **Comorbidities of the LC Cohort** | | | |
| Condition | | | |
| Hypercholesterolemia | 3/20 | Fibromyalgia | 1/20 |
| Hypertension | 1/20 | Pulmonary Embolism | 1/20 |
| Cancer | 1/20 | Psoriasis | 1/20 |
| Hypothroidism | 1/20 | Hypotension | 1/20 |
| Rosacea | 1/20 | Gingivitis | 1/20 |
| **Symptoms of the LC Cohort** | | | |
| Persistent Fatigue | 18/20 | Perturbed Sleep | 12/20 |
| Brain Fog | 16/20 | Palpitations | 11/20 |
| Post-Exertional Malaise | 13/20 | Depression | 11/20 |
| Dyspnea | 13/20 | Gastrointesinal Issues | 10/20 |
| Myalgia | 12/20 | Chest Pains | 9/10 |

**Figure S1:** Age distributions between the control and Long COVID groups. Data are shown as median [IQR]. Statistical significance was determined at p < 0.05. Significance values: * = p < 0.05, ** = p < 0.01, *** = p < 0.001, and **** = p < 0.0001.

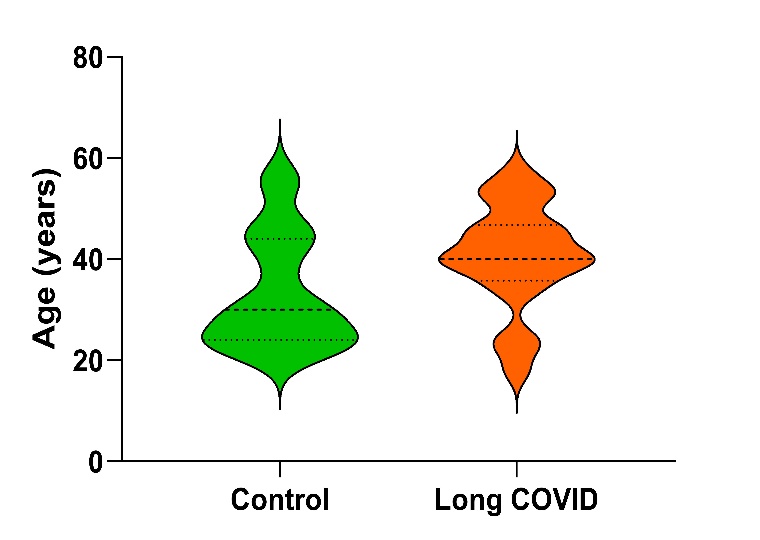

**Thromboelastography^®^ Analysis of Whole-Blood and Platelet-Poor Plasma**

Whole-blood was gently inverted prior to being analysed. For each sample, 340μL of whole-blood or PPP was pipetted into a TEG^®^ cup. This was followed by the addition of 20μL of 0.01M calcium chloride to initiate coagulation. The test was considered complete when a maximum amplitude of clot formation was achieved. The parameters assessed include Reaction time (R), Kinetics (K), α-angle, maximum amplitude (MA), maximum rate to thrombus generation (MRTG), time to maximum rate of thrombus generation (TMRTG), and total thrombus generation (TTG). The TEG^®^ was calibrated according to the manufacturer's settings, and an e-test was run prior to sample analysis.

**Imaging Flow Cytometry: Gating and Masking Strategies**

Masking of PPP events were conducted using an erosion-based mask applied to the brightfield channel (channel 01), in order to accurately determine size of events (Figure 2). There are inherent challenges in applying masks to events spanning a wide size range and exhibiting heterogeneous morphologies, as is the case in PPP, because a single mask may not accurately define the boundaries of both small and large, variably shaped events. Since we are not gating based on strict morphological criteria and are instead including all events within a broad size range (1–1000 µm²), the masking strategy does not result in exclusion of events from the analysis gate.

Since FMCs (and other objects in PPP) range from submicron to 400µm^2^ and above, the acquisition and analyses gates used have a large area range (Figure 3), which is atypical for cell-based flow cytometry where target populations often reside in exclusive gates with a narrow area range. Similar gating strategies were used for each ThT-, CD62P antibody-, and Hoechst-stained samples, with the exception of variations in masking values. Events from different areas within an exclusion gate is shown in Figure 4.

**Figure S2:** Masking strategy used for PPP events in flow cytometry experiments. Samples stained with either ThT, CD62P, or Hoechst all received an erode mask. A) ThT-stained samples (Erode(Ch01, 2)), B) CD62P antibody-stained samples (Erode(Ch01, 4)) and C) Hoechst-stained samples (Erode(Ch01, 4)). All images were taken at 20X magnification.

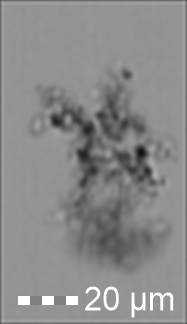

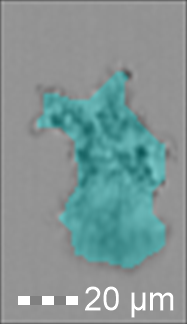

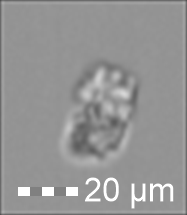

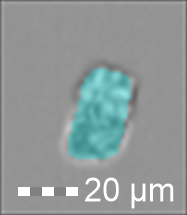

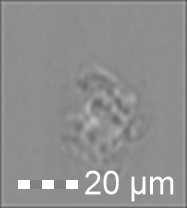

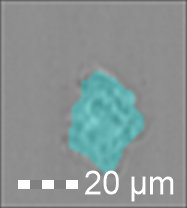

**A**

**B**

**C**

**Figure S3:** Gating strategy used for aggregates, including FMCs and cellular material, in PPP samples. A) Acquisition gate based on area and fluorescent intensity of events. B) Aspect ratio gate used to rid of events that do not contain any material in the brightfield image. C) Gradient RMS gate with a low-end value of 20 to exclude events that are signficantly out of focus. D) Size distribution of events/counts within given area bins, using the respective erode mask.

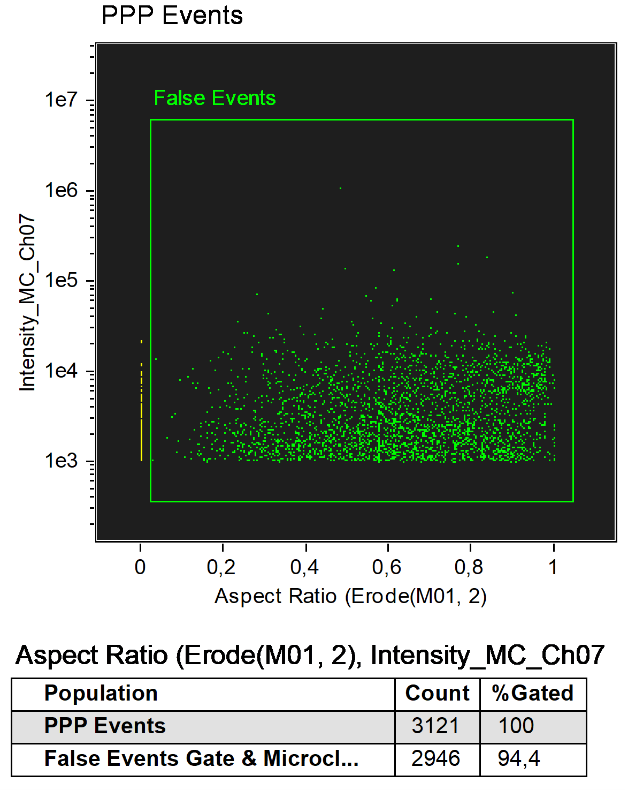

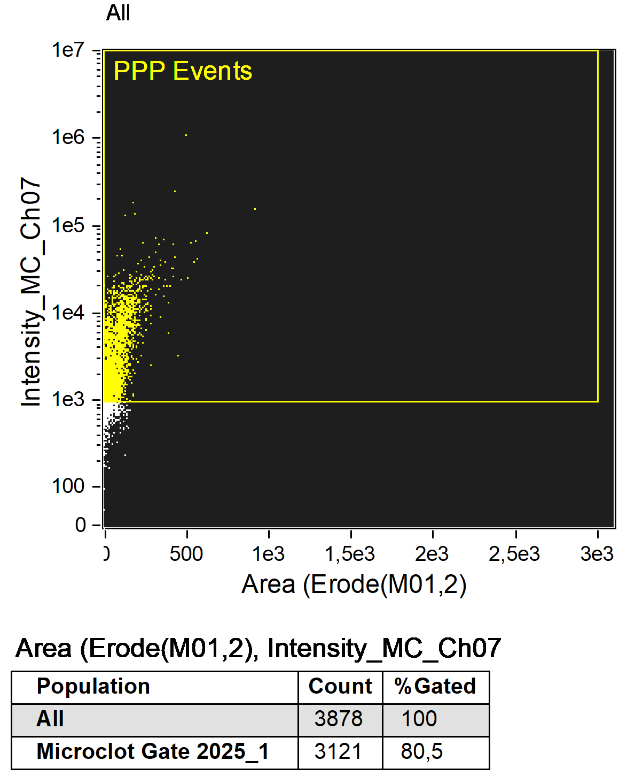

**B**

**A**

**D**

**C**

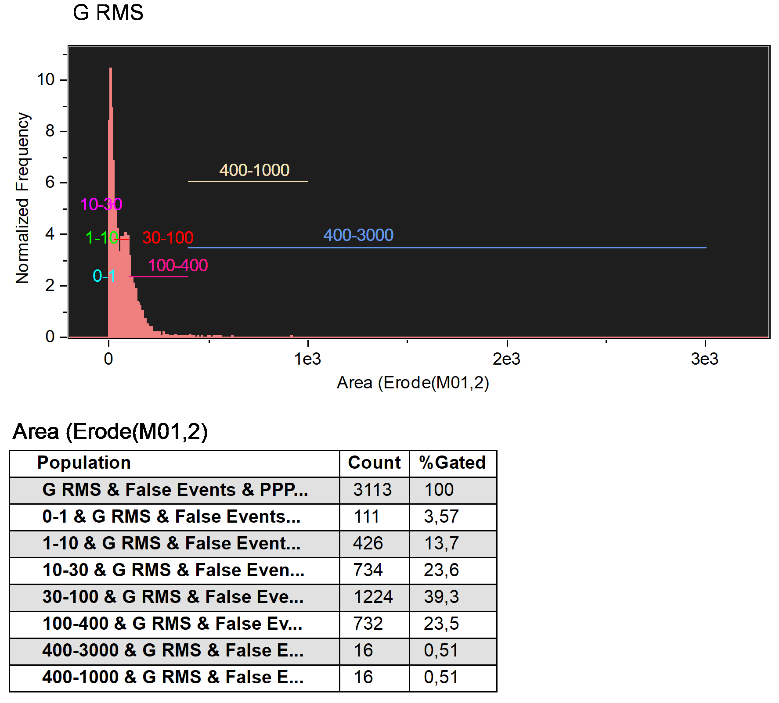

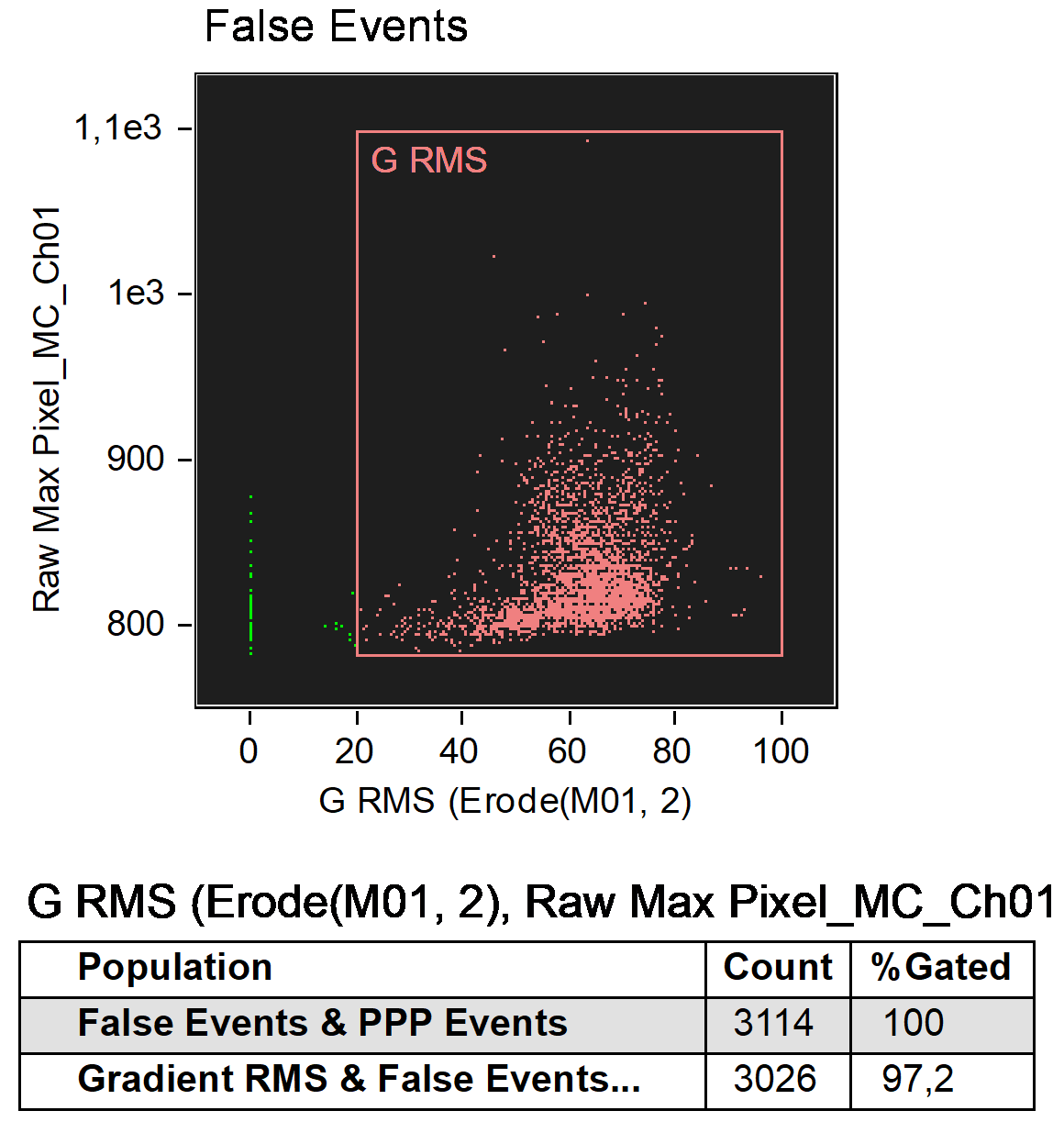

**Figure S4:** Events within an exclusion gate. This figure demonstrates how gating for events in PPP in these experiments are significantly inclusive in terms of size and morphology. An image of the event designated by A-G is presented on the right of the graph.

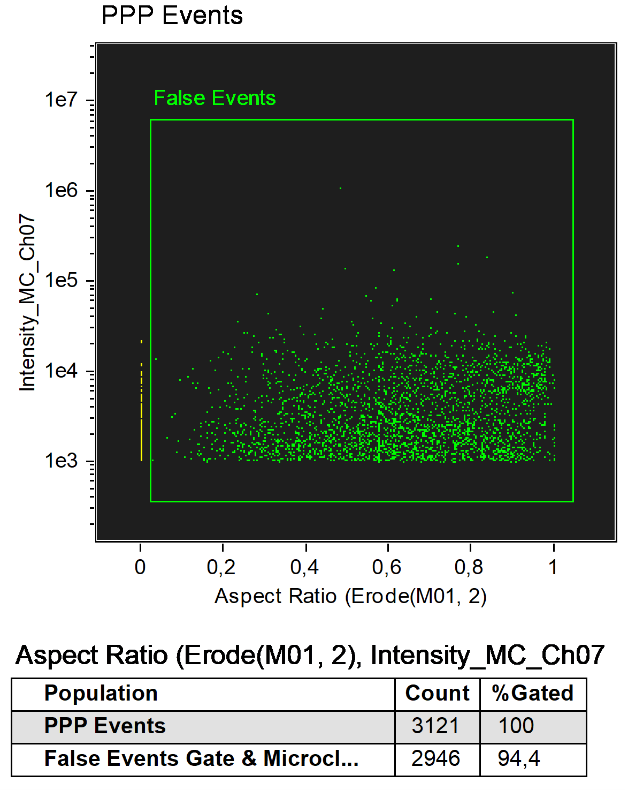

**A**

**B**

**C**

**D**

**E**

**F**

**G**

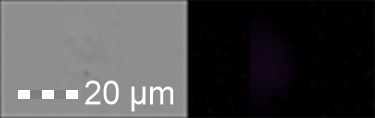

**A**

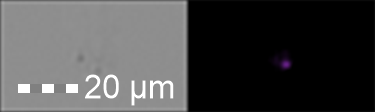

**B**

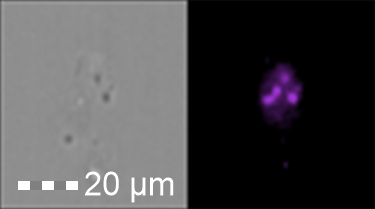

**C**

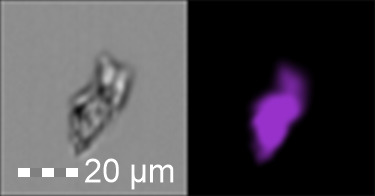

**D**

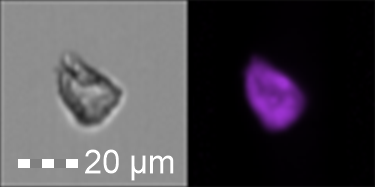

**E**

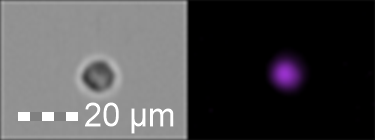

**F**

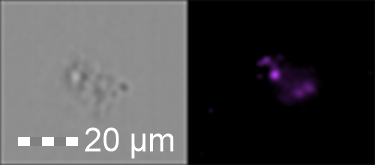

**G**

**Confocal laser scanning microscopy**

After PPP samples were thawed at room temperature, 29 μL of PPP was combined with 1 μL of ThT and 20 μL of CellMask™ Red. Staining was performed in low-light conditions and samples were incubated for 30 minutes. 5 μL of co-stained ThT and CellMask™ Red PPP was placed on a microscope slide, smeared, and a coverslip was applied. Confocal imaging was performed using an Evident Flouroview 4000 laser scanning confocal microscope using the UPlanXApo 60x/1.42 objective (Tokyo, Japan). For ThT, excitation was achieved using the 488 nm laser, and emission was collected between 500-599 nm. For CellMask™ Red, the 640 nm laser was used for excitation, and the emission range was set between 650-690 nm. In addition, brightfield images were acquired under identical optical conditions to provide additional morphological data. To account for autofluorescence, acquisition parameters were standardised across all samples to minimise variation due to detector gain. Specifically, photon detection thresholds were set to 0-50 photons for the ThT channel and 0-150 photons for the CellMask™ Red channel.

**Fluorescence microscope**

To be able assess the isolated “crescent pellet”, the PPP samples were centrifuged at 16,000 x g for 10 minutes at room temperature and then another 10 minutes at 0°C. The samples were stained for MPO (exposure concentration: 0,125 μg; 48-1299-42, Invitrogen, Waltham, MA, USA) and ThT (exposure concentration: 5 μM) and incubated for 30 min in the dark. After placing a 5 μL drop of the sample on a microscope slide, the sample was viewed with a Zeiss Axio Observer 7 fluorescence microscope with a Plan-Apochromat 63 ×/1.4 Oil DIC M27 objective (Carl Zeiss Microscopy, Munich, Germany) using the excitation wavelength of 370-400 nm and emission from 410 to 440 nm for the MPO and excitation wavelength of 450-488 nm and emission from 499 to 529 nm for the ThT.

**Polarized microscope**

3 µL of Congo Red (0.3 mM final concentration) (1.01641.0001, Sigma-Aldrich) was added to 46 µL of PPP. and 1 µL of ThT to the samples for the amyloid analysis. For the presence of platelet debris, we added 1.2 µL of Congo Red to 14.8 µL of PPP and 4 µL PAC-1 (FITC-conjugated) (340507, BD Biosciences, San Jose, CA, USA). For all the staining the samples were incubated in the dark for 30 minutes at room temperature. On the microscope slide 5 µL of sample was added together with 2.5µL thrombin (final concentration 6.7 IU/mL) to induce clot formation and enhance the imaging. For ThT and PAC-1, the excitation wavelength was set at 465-495 nm and emission to 515-555 nm. The Congo Red was evaluated using polarized light to detect amyloid. All images were captured using the Plan Fluor 40x/0.72 objective on the Nikon Eclipse E400 microscope (Tokyo, Japan).

**Scanning Electron Microscopy (SEM)**

Whole-blood samples were prepared for SEM analysis on the same day of collection to preserve cellular and structural integrity. 5μL of whole-blood was carefully placed onto cover slip discs contained within a 24-well plate. The samples were allowed to air-dry for 1 minute. PBS was then gently introduced along the wall of each well to remove excess plasma and unbound elements while minimising direct disturbance of the sample. The samples were then fixed using 4% formaldehyde for 30 minutes. Following fixation with formaldehyde, samples were washed with PBS three times for 3 minutes each to remove residual fixative, and were then fixed in 1% osmium tetroxide for 15 minutes, which also enhances membrane contrast and electron density. This was followed by three more wash steps.

Dehydration was achieved through a graded ethanol exposure for 3 minutes each, namely 30%, 50%, 70%, 90%, and 100%. Samples were then immersed in HMDS for 30 minutes to facilitate drying and minimise surface tension artefacts. Following HMDS treatment, the 24-well plate was left in a biosafety cabinet with the lid removed and allowed to air-dry overnight under sterile conditions. Following sample preparation and complete air-drying, all discs were coated with carbon using a high-vacuum carbon coater. The coated samples were then examined using a Zeiss Merlin scanning electron microscope (Carl Zeiss, Germany). Micrographs were obtained using the InLens at 1kV. Representative micrographs were captured for both control and Long COVID groups.

**Persistent Hypercoagulability and the Distinctions Between and Associations of FMCs and Cellular Material in Long COVID**

The following paragraphs further clarifies the statistical analyses of the imaging flow-cytometry data that are referenced but not reported in full in the main text. It covers the five staining experiments (undiluted PPP + ThT, 10× diluted PPP + ThT, and the resuspended PPP pellet stained separately with ThT, CD62P‑PE, or Hoechst) for the control (n = 19) and Long COVID (n = 20) cohorts. All analyses were performed in R (v4.5.2); package versions are given in the main-text Statistical Analysis section.

Two independent aspects of the data are reported separately throughout: abundance (how many objects were detected: event counts) and composition (the shape of the size distribution: within-sample proportions across the six area bins, independent of abundance). The six area bins are 0–1, 1–10, 10–30, 30–100, 100–400 and 400–3000 µm². Significance symbols: ns q ≥ 0.05, * q < 0.05, ** q < 0.01, *** q < 0.001.

**Abundance: full per-bin statistics**

Control vs Long COVID were compared with the two-sided Mann–Whitney U test (primary) and a Welch *t*-test (parametric cross-check). The total ("All") is a single primary endpoint per experiment and is reported unadjusted; the six size bins within each experiment form one Benjamini–Hochberg (BH) family, reported as *q*. Values are group medians; *p* is the Mann–Whitney *p*, *q* the BH-adjusted value across the six bins, and *p*(Welch) the parametric cross-check.

**Table S2.** Event counts, Control vs Long COVID, per experiment × size bin.

| Experiment | Bin | Median (Control) | Median (LC) | *p* (MW) | *q* (BH) | sig (*q*) | *p* (Welch) |
| --- | --- | --- | --- | --- | --- | --- | --- |
| Undiluted ThT | **All** | 678 | 1766 | 0.019 | n/a | ***** | 0.477 |
| Undiluted ThT | 0–1 | 39 | 90 | 0.021 | 0.032 | ***** | 0.210 |
| Undiluted ThT | 1–10 | 67 | 305 | 0.012 | 0.032 | ***** | 0.085 |
| Undiluted ThT | 10–30 | 106 | 534 | 0.006 | 0.032 | ***** | 0.027 |
| Undiluted ThT | 30–100 | 201 | 621 | 0.019 | 0.032 | ***** | 0.696 |
| Undiluted ThT | 100–400 | 230 | 441 | 0.100 | 0.120 | ns | 0.454 |
| Undiluted ThT | 400–3000 | 6 | 20 | 0.254 | 0.254 | ns | 0.831 |
| Diluted ThT | **All** | 600 | 990 | 0.0024 | n/a | ****** | 0.028 |
| Diluted ThT | 0–1 | 3 | 5 | 0.164 | 0.164 | ns | 0.184 |
| Diluted ThT | 1–10 | 7 | 12 | 0.053 | 0.088 | ns | 0.125 |
| Diluted ThT | 10–30 | 7 | 14.5 | 0.073 | 0.088 | ns | 0.102 |
| Diluted ThT | 30–100 | 12 | 33 | 3.6e−05 | 0.0002 | ******* | 0.003 |
| Diluted ThT | 100–400 | 18 | 35.5 | 0.039 | 0.088 | ns | 0.163 |
| Diluted ThT | 400–3000 | 2 | 4.5 | 0.066 | 0.088 | ns | 0.107 |
| Pellet ThT | **All** | 281 | 527 | 0.032 | n/a | ***** | 0.021 |
| Pellet ThT | 0–1 | 7 | 17 | 0.221 | 0.265 | ns | 0.046 |
| Pellet ThT | 1–10 | 21 | 33 | 0.129 | 0.194 | ns | 0.038 |
| Pellet ThT | 10–30 | 40 | 75 | 0.079 | 0.194 | ns | 0.098 |
| Pellet ThT | 30–100 | 87 | 149.5 | 0.046 | 0.194 | ns | 0.045 |
| Pellet ThT | 100–400 | 96 | 153.5 | 0.097 | 0.194 | ns | 0.047 |
| Pellet ThT | 400–3000 | 6 | 7 | 0.353 | 0.353 | ns | 0.049 |
| Pellet CD62P | **All** | 7718 | 14761 | 0.0021 | n/a | ****** | 0.004 |
| Pellet CD62P | 0–1 | 102 | 38 | 0.989 | 0.989 | ns | 0.063 |
| Pellet CD62P | 1–10 | 170 | 332.5 | 0.005 | 0.016 | ***** | 0.049 |
| Pellet CD62P | 10–30 | 1696 | 2907.5 | 0.056 | 0.083 | ns | 0.044 |
| Pellet CD62P | 30–100 | 4646 | 6452 | 0.050 | 0.083 | ns | 0.091 |
| Pellet CD62P | 100–400 | 131 | 281.5 | 0.0024 | 0.014 | ***** | 0.022 |
| Pellet CD62P | 400–3000 | 4 | 12 | 0.069 | 0.083 | ns | 0.030 |
| Pellet Hoechst | **All** | 235419 | 162378 | 0.687 | n/a | ns | 0.531 |
| Pellet Hoechst | 0–1 | 27404 | 16736 | 0.120 | 0.701 | ns | 0.230 |
| Pellet Hoechst | 1–10 | 86884 | 67475 | 0.247 | 0.701 | ns | 0.190 |
| Pellet Hoechst | 10–30 | 65758 | 43742 | 0.513 | 0.701 | ns | 0.489 |
| Pellet Hoechst | 30–100 | 38810 | 28462 | 0.411 | 0.701 | ns | 0.806 |
| Pellet Hoechst | 100–400 | 14060 | 20297 | 0.901 | 0.901 | ns | 0.328 |
| Pellet Hoechst | 400–3000 | 1107 | 1344 | 0.584 | 0.701 | ns | 0.074 |

The four elevated totals (both ThT preparations, Pellet ThT, CD62P) remain significant if the five "All" totals are themselves BH-corrected across experiments (all *q* < 0.04); Hoechst remains non-significant. The elevation is broad across the size spectrum: 4/6 undiluted-ThT bins and several diluted-ThT and CD62P bins survive FDR.

**Dispersion: is the greater spread in Long COVID an independent increase in heterogeneity?**

Absolute counts are more variable in Long COVID, but this reflects mean–variance coupling (variance scaling with the higher counts) rather than an independent increase in relative heterogeneity. Homogeneity of variance on the total count ("All") was tested with the Fligner–Killeen test on both the raw and the log₁₀-transformed counts and summarised with two scale-free measures: the coefficient of variation (CV) and the log-scale SD, each expressed as a Long COVID ÷ Control ratio.

**Table S3.** Dispersion of the total count ("All"), Control vs Long COVID.

| Experiment | Fligner *p* (raw) | Fligner *p* (log₁₀) | CV (Ctrl) | CV (LC) | CV ratio | logSD (Ctrl) | logSD (LC) | logSD ratio |
| --- | --- | --- | --- | --- | --- | --- | --- | --- |
| Undiluted ThT | 0.134 | 0.792 | 2.37 | 1.68 | 0.71 | 0.60 | 0.51 | 0.85 |
| Diluted ThT | 0.059 | 0.762 | 0.66 | 1.18 | 1.80 | 0.30 | 0.34 | 1.15 |
| Pellet ThT | **0.001** | 0.495 | 1.38 | 1.38 | 1.00 | 0.68 | 0.75 | 1.11 |
| Pellet CD62P | 0.060 | 0.673 | 0.80 | 0.80 | 1.00 | 0.41 | 0.35 | 0.85 |
| Pellet Hoechst | 0.652 | 0.359 | 0.70 | 0.83 | 1.19 | 0.52 | 0.56 | 1.07 |

On the log scale the between-group variance difference is non-significant for every experiment (Fligner–Killeen, all *p* ≥ 0.36), and the log-scale SD ratios sit near 1 (0.85–1.15). The one experiment with a raw-scale variance difference (Pellet ThT, *p* = 0.001) loses it after log-transformation, the signature of mean–variance coupling. The CV ratios are near unity except for Diluted ThT (1.80); the log-scale SD is the more stable scale-free summary here and is near
unity throughout.

The whole-distribution view is shown in **Fig. S1**: the Long COVID − Control shift function (Harrell–Davis deciles, 95% percentile-bootstrap CI, 2000 resamples; top row) and the Long COVID ÷ Control quantile ratio (bottom row). The ratio is approximately flat near a constant multiplier for undiluted-ThT and CD62P (consistent with a multiplicative up-shift rather than a widening); it is noisier for the pellet preparations, where the upper-tail ratio rises (Table S3), so the robust support for the "no extra heterogeneity" conclusion is the log-scale Fligner–Killeen and log-SD result above rather than the quantile-ratio curve alone. **Fig. S2** (output/plots/lev/lev_spread_location.png) plots SD against median (log–log) for every experiment × size bin: both groups fall on a common line, i.e. spread is set by the level, not by group.

**Figure S5:** Total count across the distribution: the absolute gap LC – Control (top) and LC/Control ratio (bottom)

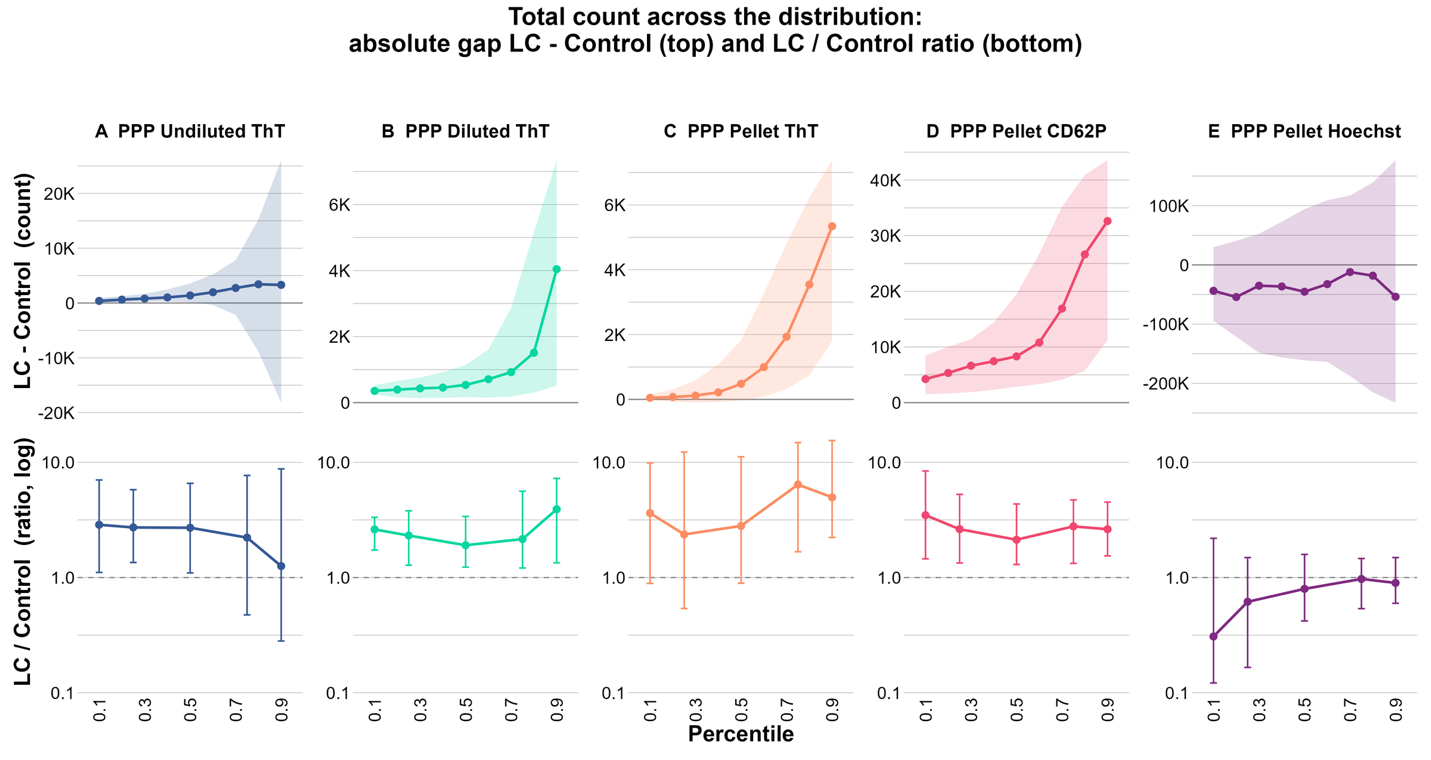

**Figure S6:** Spread location. LC variance tracks its level.

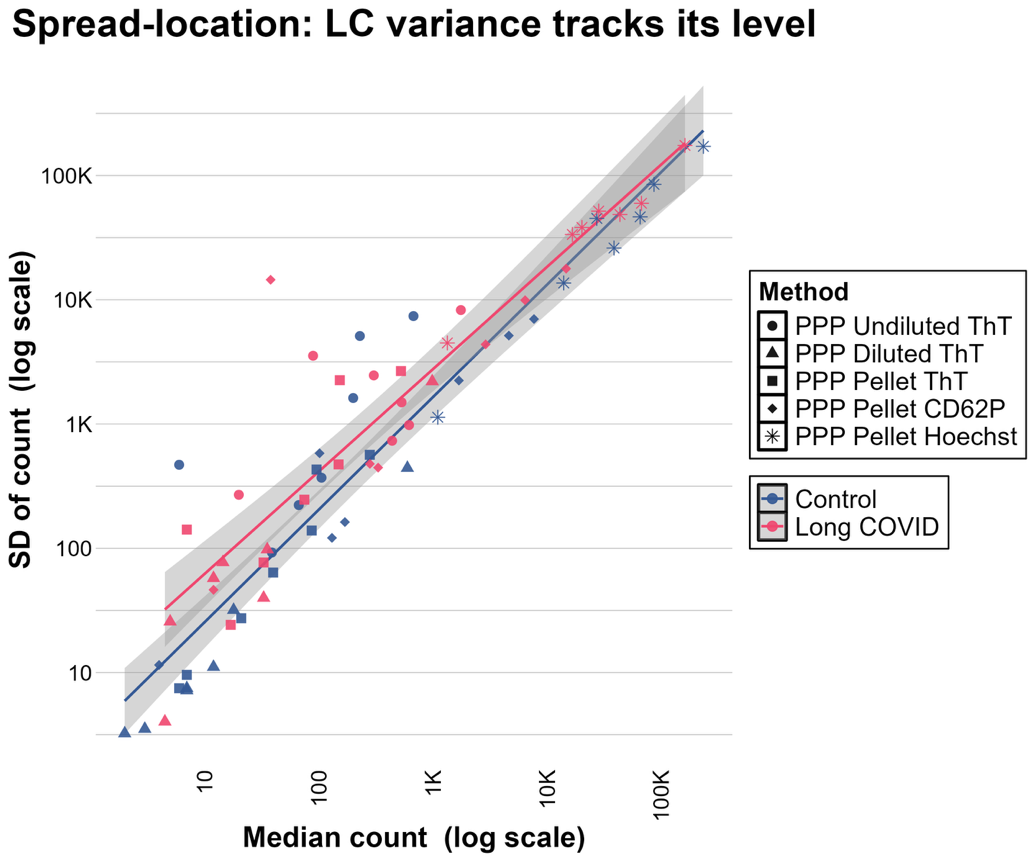

**Table S4.** Long COVID ÷ Control quantile ratio of the total count (Harrell–Davis estimator).

| Experiment | p10 | p25 | p50 | p75 | p90 |
| --- | --- | --- | --- | --- | --- |
| Undiluted ThT | 2.87 | 2.72 | 2.71 | 2.22 | 1.26 |
| Diluted ThT | 2.62 | 2.32 | 1.91 | 2.16 | 3.91 |
| Pellet ThT | 3.63 | 2.37 | 2.80 | 6.41 | 4.97 |
| Pellet CD62P | 3.48 | 2.63 | 2.13 | 2.78 | 2.63 |
| Pellet Hoechst | 0.31 | 0.62 | 0.80 | 0.97 | 0.90 |

**Correlation of FMC counts across paired PPP preparations**

The three ThT preparations (undiluted, diluted, resuspended pellet) were performed on the same 39 participants, so FMC recovery can be compared across preparations within participant. Spearman rank correlations of the total ("All") counts are reported overall and within each cohort.

**Table S5.** Spearman correlation (ρ, *p*) of ThT total counts between preparations.

| Preparations | Overall (n=39) | Control (n=19) | Long COVID (n=20) |
| --- | --- | --- | --- |
| Undiluted vs Diluted | ρ = 0.17, *p* = 0.29 | ρ = −0.17, *p* = 0.49 | ρ = 0.19, *p* = 0.43 |
| Undiluted vs Pellet | ρ = −0.02, *p* = 0.89 | ρ = 0.09, *p* = 0.72 | ρ = −0.43, *p* = 0.063 |
| Diluted vs Pellet | ρ = −0.16, *p* = 0.33 | ρ = −0.13, *p* = 0.58 | **ρ = −0.62, *p* = 0.003** |

Across the full cohort, and within controls, ThT counts are **not positively** **correlated** across preparations: no participant-level concordance in FMC recovery, consistent with substantial sample-to-sample variability introduced by dilution and pelleting. Within Long COVID there is one significant association: Diluted vs Pellet counts are **negatively** correlated (ρ = −0.62, *p* = 0.003; Kendall τ = −0.49, *p* = 0.003; Pearson on log₁₀ = −0.49, *p* = 0.029). It is robust to leave-one-out resampling (*p* ≤ 0.011 with any single participant removed) and survives BH correction across the three Long COVID pairs (*q* ≈ 0.01). A negative rather than positive association reinforces, rather than contradicts, the absence of concordant recovery: participants with the most FMCs recovered in the diluted preparation tended to yield the fewest in the pellet.

**Size composition: the shape of the size distribution**

Each sample's six bin counts were converted to within-sample proportions (each bin ÷ the sum of the six bins), removing sample-to-sample differences in total abundance and isolating the shape of the size distribution.

***Per-bin proportion comparisons***

Within-sample bin proportions were compared between groups with the Mann–Whitney U test, BH-corrected across the six bins per experiment. Per-experiment distributions are shown in **Fig. S3.A–E**

**Figure S7.A**

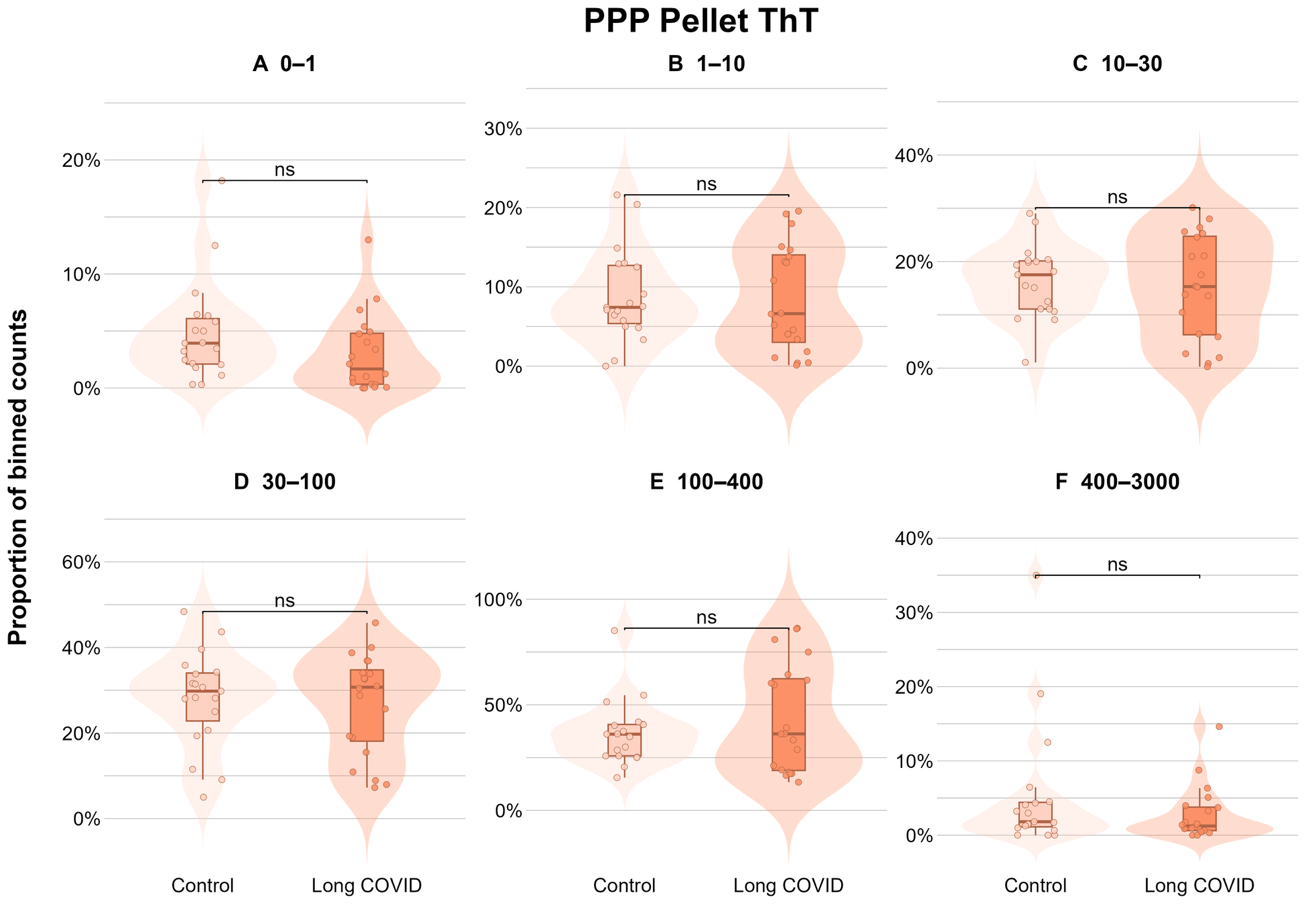

**Figure S7.B**

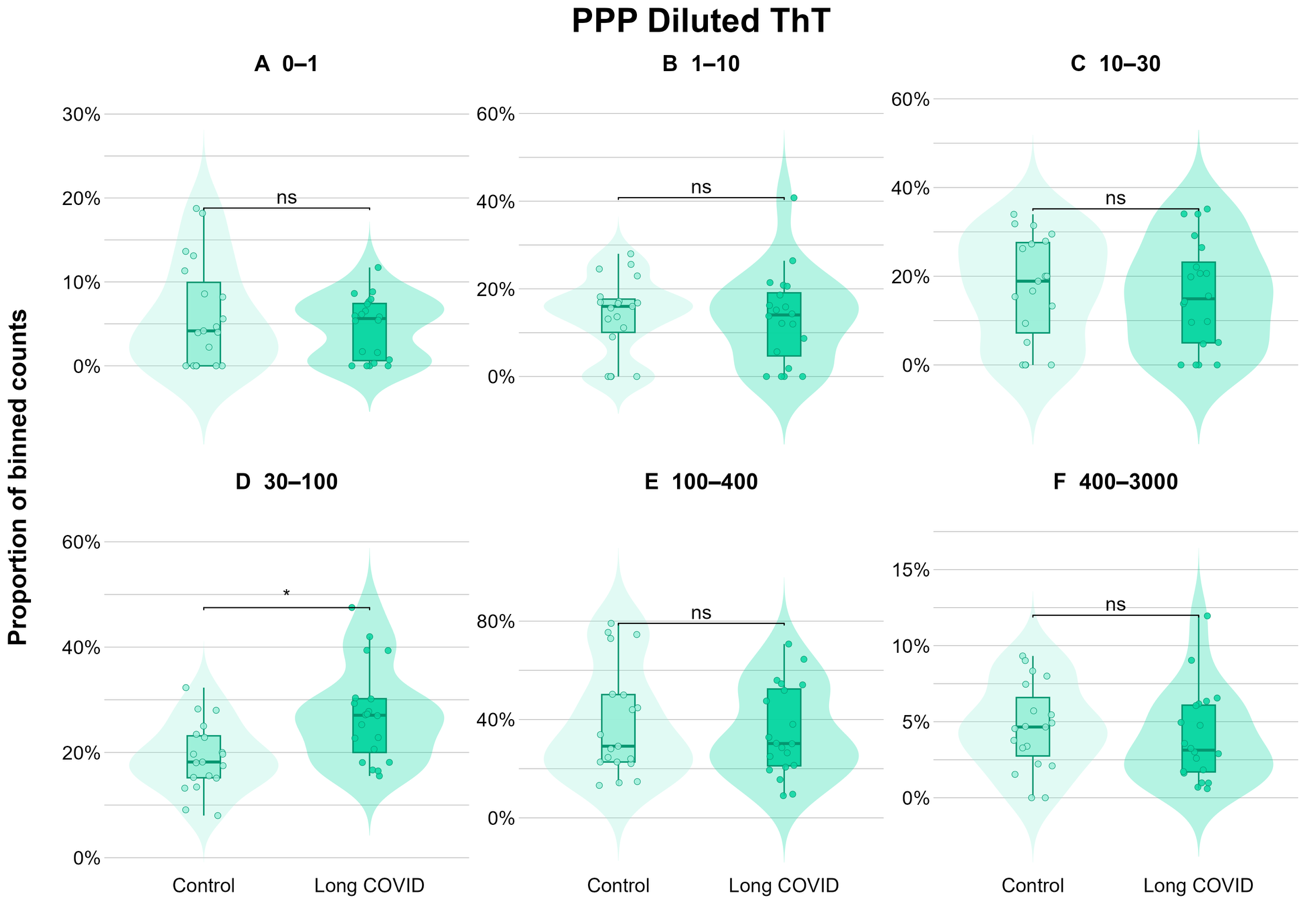

**Figure S7.C**

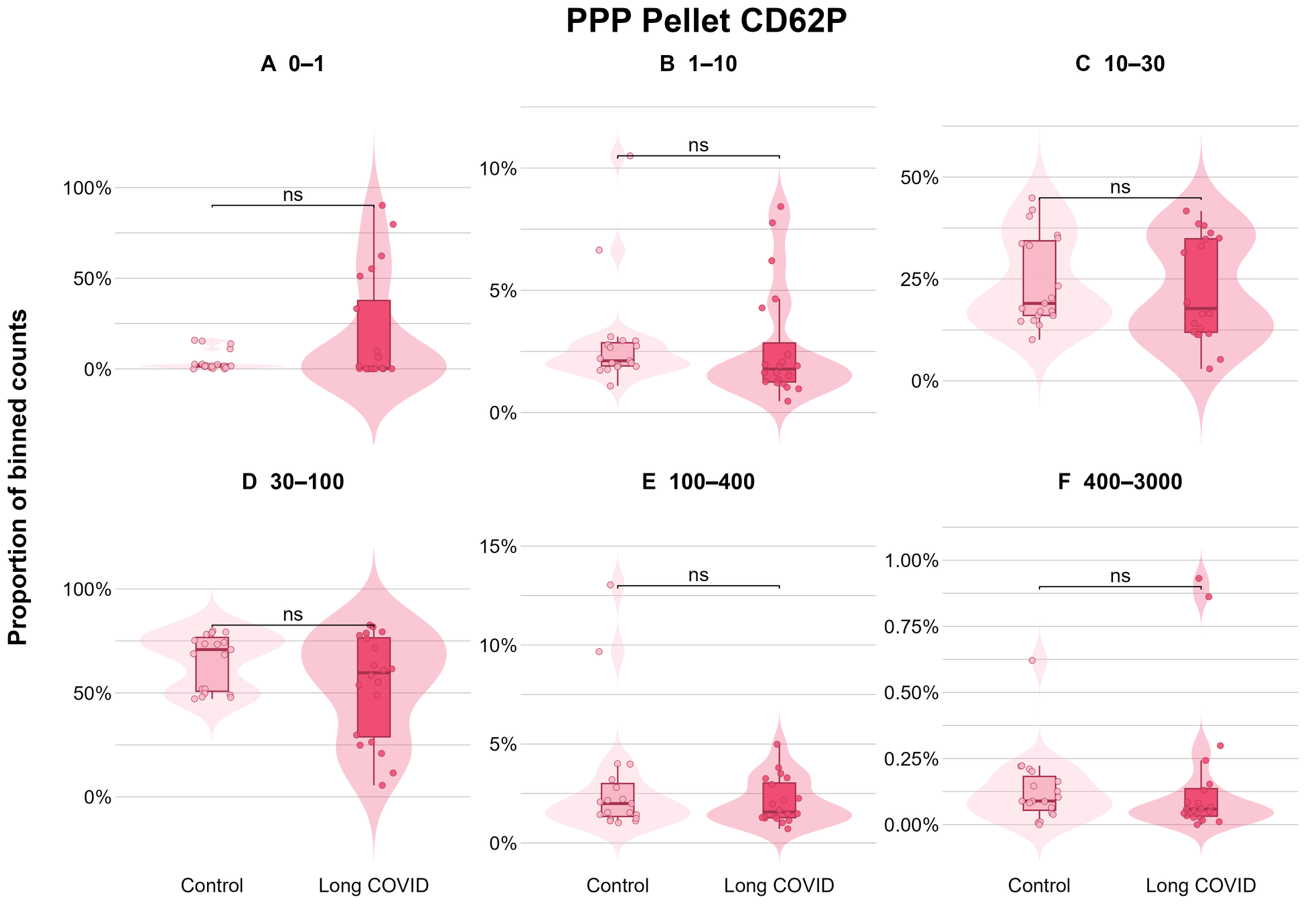

**Figure S7.D**

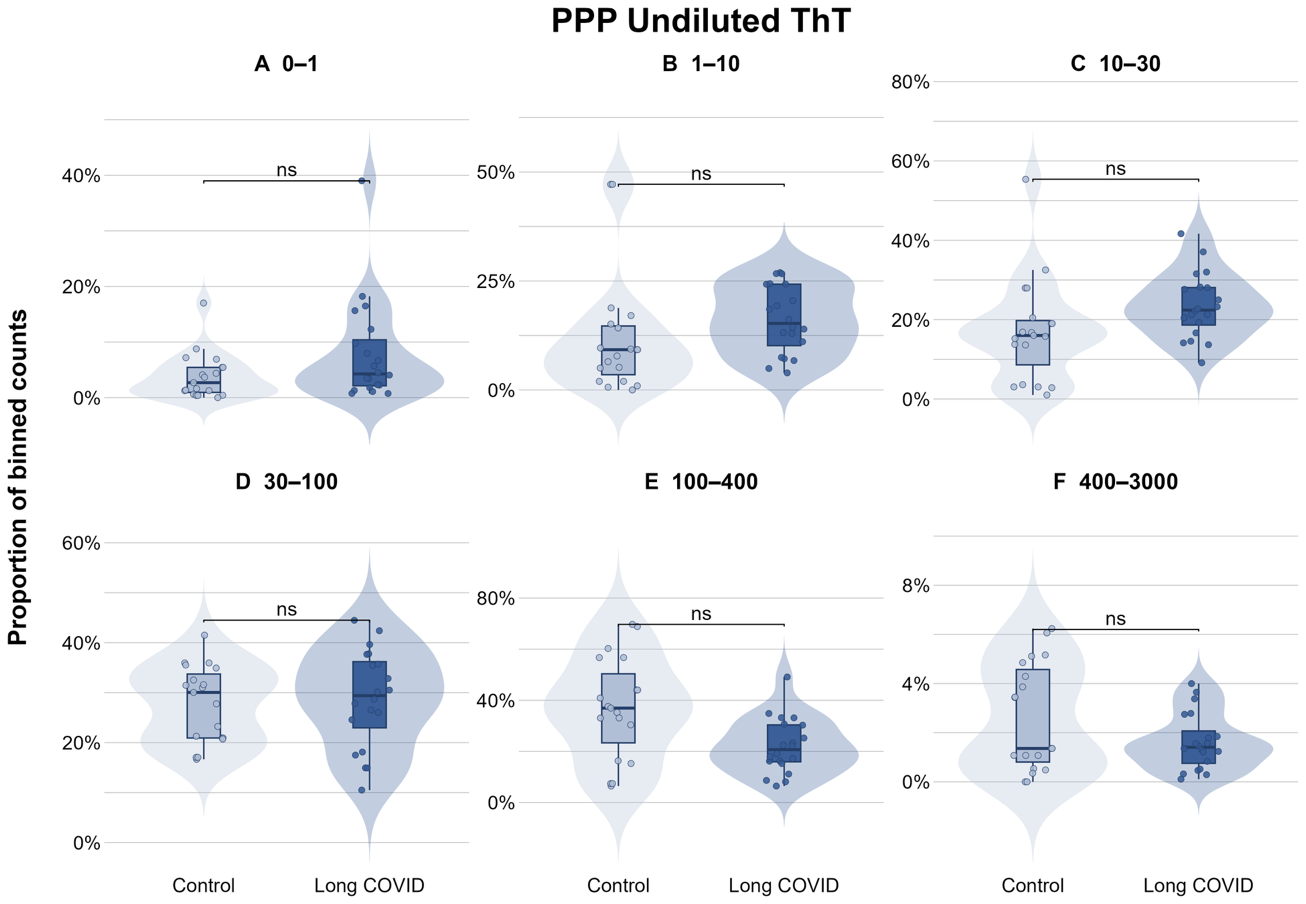

**Figure S7.E**

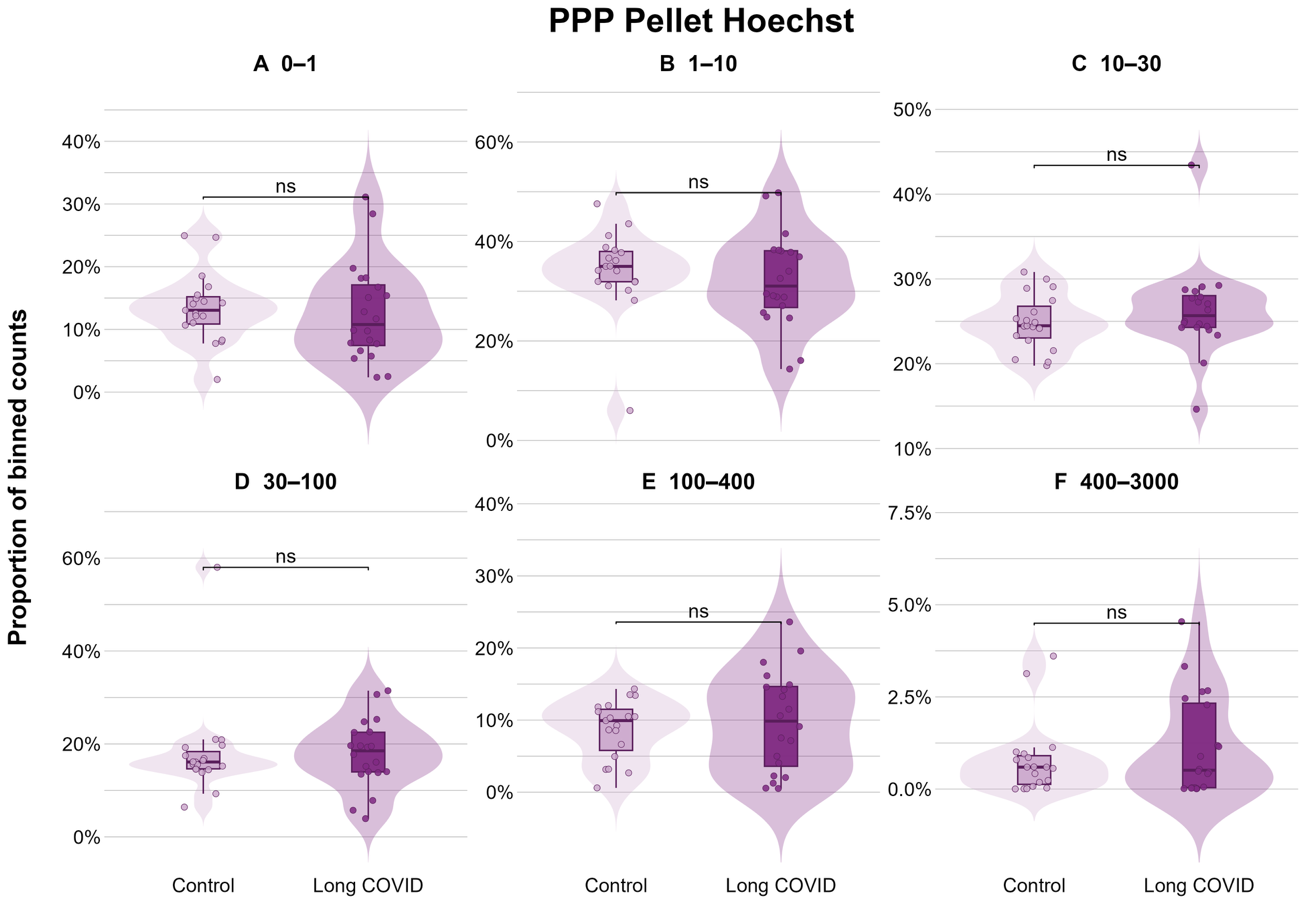

**Table S6.** Mann–Whitney comparison of within-sample bin proportions (*p*, and BH *q* across the six bins).

| Experiment | Bin | *p* | *q* | sig (*q*) |
| --- | --- | --- | --- | --- |
| Undiluted ThT | 0–1 | 0.095 | 0.142 | ns |
| Undiluted ThT | 1–10 | 0.032 | 0.063 | ns |
| Undiluted ThT | 10–30 | 0.022 | 0.063 | ns |
| Undiluted ThT | 30–100 | 0.684 | 0.684 | ns |
| Undiluted ThT | 100–400 | 0.016 | 0.063 | ns |
| Undiluted ThT | 400–3000 | 0.332 | 0.398 | ns |
| Diluted ThT | 30–100 | 0.0067 | 0.040 | ***** |
| Diluted ThT | (0–1, 1–10, 10–30, 100–400, 400–3000) | 0.41–0.90 | 0.90 | ns |
| Pellet ThT | (all six bins) | 0.079–0.95 | 0.47–0.95 | ns |
| Pellet CD62P | (all six bins) | 0.094–0.71 | 0.55–0.71 | ns |
| Pellet Hoechst | (all six bins) | 0.32–0.67 | 0.67 | ns |

Only one per-bin proportion difference survives correction (Diluted ThT, 30–100 µm², *q* = 0.040); in undiluted ThT the 1–10 and 10–30 bins are raised and 100–400 is lowered in Long COVID at the uncorrected level (all *q* ≈ 0.063), the coherent "shift to smaller" signal examined further below.

***Size-shift index***

Each sample's distribution was summarised as the count-weighted mean size-bin rank (bins ranked 1 = smallest to 6 = largest); higher = distribution shifted toward larger objects. Groups were compared with the Mann–Whitney U test, BH-corrected across the five experiments (**main-text Fig. 4A**).

**Table S7.** Size-shift index (count-weighted mean bin rank), median per group.

| Experiment | Median (Control) | Median (LC) | *p* | *q* | sig (*q*) |
| --- | --- | --- | --- | --- | --- |
| Undiluted ThT | 3.93 | 3.45 | 0.036 | 0.182 | ns |
| Diluted ThT | 3.83 | 3.69 | 0.923 | 0.923 | ns |
| Pellet ThT | 4.03 | 4.01 | 0.857 | 0.923 | ns |
| Pellet CD62P | 3.70 | 3.61 | 0.134 | 0.335 | ns |
| Pellet Hoechst | 2.75 | 2.84 | 0.550 | 0.917 | ns |

No experiment shows a size-index shift after correction.

***Multivariate composition: PERMANOVA and PERMDISP***

The six-bin proportion vector of each sample was treated as a composition. Exact zeros (~4% of entries) received a pseudocount of 0.5 before the centred log-ratio (CLR) transform; the Aitchison distance is the Euclidean distance between CLR vectors. Per experiment, PERMANOVA (adonis2, 9999 permutations) tested for a difference in mean composition and PERMDISP (betadisper, 9999 permutations) for a difference in compositional dispersion. Ordination is shown in **main-text** **Fig. 5**.

**Table S8.** PERMANOVA and PERMDISP per experiment.

| Experiment | PERMANOVA R² | PERMANOVA *p* | PERMDISP *p* | Dispersion (Ctrl) | Dispersion (LC) | More dispersed |
| --- | --- | --- | --- | --- | --- | --- |
| Undiluted ThT | 0.080 | **0.039** | 0.085 | 1.96 | 1.38 | Control (trend) |
| Diluted ThT | 0.014 | 0.540 | 0.941 | 1.85 | 1.89 | n/a |
| Pellet ThT | 0.028 | 0.334 | 0.173 | 1.61 | 2.07 | n/a |
| Pellet CD62P | 0.005 | 0.818 | **0.009** | 1.71 | 3.23 | Long COVID |
| Pellet Hoechst | 0.005 | 0.808 | 0.323 | 1.54 | 1.97 | n/a |

Two experiments depart from equivalence. **Undiluted ThT** shows a modest between-group composition difference (PERMANOVA *p* = 0.039); because PERMDISP also trends (control more dispersed, *p* = 0.085) this is partly dispersion-driven and is described as a *suggestive* shift toward smaller events. **Pellet CD62P** shows no centroid difference (PERMANOVA *p* = 0.82) but markedly greater dispersion in Long COVID (PERMDISP *p* = 0.009; dispersion 1.71 → 3.23), a compositional counterpart to the elevated and more variable activated-platelet counts.

**Equivalence of the size-shift index**

To distinguish "no evidence of a difference" from "evidence of no meaningful difference," the between-group shift in the size-shift index was estimated with the Hodges–Lehmann estimator (median of all pairwise Control − Long COVID differences) and its 95% CI and evaluated against candidate equivalence margins. Equivalence holds when the CI lies entirely within the margin. Because no field-standard margin exists, the verdict was swept across ±0.3, ±0.5, ±0.75 and ±1.0 bins (**main-text** **Fig. 4B**).

**Table S9.** Hodges–Lehmann shift (Control − Long COVID) and equivalence verdict
across margins.

| Experiment | HL shift | 95% CI | ±0.3 | ±0.5 | ±0.75 | ±1.0 |
| --- | --- | --- | --- | --- | --- | --- |
| Undiluted ThT | 0.484 | [0.025, 0.831] | ✗ | ✗ | ✗ | ✓ |
| Diluted ThT | −0.031 | [−0.495, 0.337] | ✗ | ✓ | ✓ | ✓ |
| Pellet ThT | −0.041 | [−0.440, 0.284] | ✗ | ✓ | ✓ | ✓ |
| Pellet CD62P | 0.119 | [−0.058, 0.504] | ✗ | ✗ | ✓ | ✓ |
| Pellet Hoechst | −0.096 | [−0.344, 0.172] | ✗ | ✓ | ✓ | ✓ |

Diluted ThT, Pellet ThT and Hoechst are equivalent from ±0.5 upward (robust); CD62P only from ±0.75 (the wide-but-unshifted profile); Undiluted ThT not until ±1.0 (the genuine shift). At the strict ±0.3 margin no experiment is equivalent, reflecting limited power at n ≈ 20; equivalence is therefore claimed at the half-bin (±0.5) primary margin, not tighter.

***Per-bin effect size (Cliff's delta)***

For each size bin, Cliff's delta on the within-sample proportions was computed as P(LC > Control) − P(Control > LC); the sign indicates which group has the higher proportion in that bin (positive = higher in Long COVID). 95% CIs are from 2000 bootstrap resamples (**Fig. S4**).

**Figure S8:** Per-bin effect size (Cliff’s delta, 95% bootstrap CI)

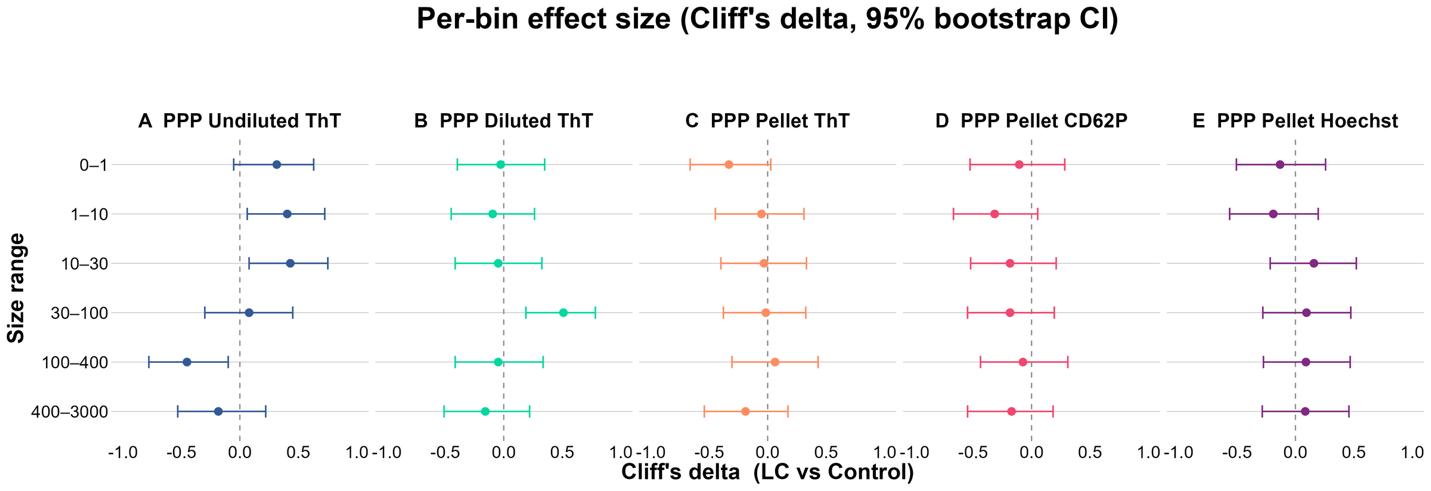

**Table S10.** Cliff's delta (Long COVID vs Control) per bin, with 95% bootstrap CI.
Bold = CI excludes 0.

| Experiment | 0–1 | 1–10 | 10–30 | 30–100 | 100–400 | 400–3000 |
| --- | --- | --- | --- | --- | --- | --- |
| Undiluted ThT | 0.32 [−0.05, 0.63] | **0.41 [0.06, 0.73]** | **0.43 [0.08, 0.75]** | 0.08 [−0.30, 0.45] | **−0.45 [−0.78, −0.10]** | −0.18 [−0.53, 0.22] |
| Diluted ThT | −0.03 [−0.40, 0.35] | −0.09 [−0.45, 0.26] | −0.05 [−0.42, 0.33] | **0.51 [0.19, 0.78]** | −0.05 [−0.42, 0.34] | −0.16 [−0.51, 0.22] |
| Pellet ThT | −0.33 [−0.66, 0.03] | −0.05 [−0.45, 0.31] | −0.03 [−0.40, 0.33] | −0.02 [−0.38, 0.33] | 0.06 [−0.31, 0.43] | −0.19 [−0.54, 0.17] |
| Pellet CD62P | −0.11 [−0.53, 0.28] | −0.32 [−0.67, 0.05] | −0.18 [−0.52, 0.21] | −0.18 [−0.55, 0.19] | −0.07 [−0.44, 0.31] | −0.17 [−0.55, 0.18] |
| Pellet Hoechst | −0.13 [−0.51, 0.26] | −0.19 [−0.56, 0.19] | 0.16 [−0.22, 0.52] | 0.09 [−0.28, 0.47] | 0.09 [−0.27, 0.47] | 0.08 [−0.28, 0.46] |

The only intervals excluding zero are in undiluted ThT (positive in 1–10 and 10–30, negative in 100–400, a coherent shift toward smaller events in Long COVID) and diluted ThT (positive at 30–100), matching Tables S5–S7.

**Large-vs-small compositional balance**

As a targeted large-vs-small contrast, a CLR log-ratio balance was computed per sample as ln(geometric-mean proportion of the three larger bins ÷ that of the three smaller bins); higher = relatively more large objects. Groups were compared with the Mann–Whitney U test, BH-corrected across the five experiments (**Fig. S5**).

**Figure S9: Large-vs-small balance: relative dominance of large clots**

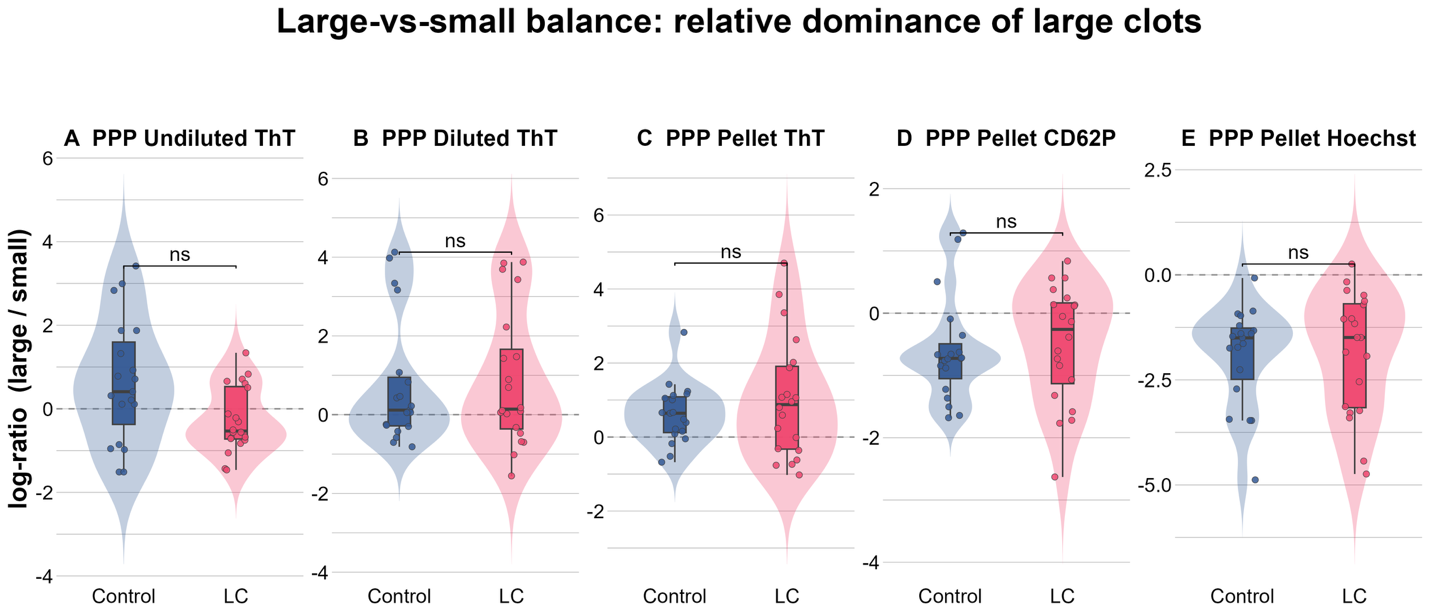

**Table S11.** Large-vs-small balance, median per group.

| Experiment | Median (Control) | Median (LC) | *p* | *q* | Direction |
| --- | --- | --- | --- | --- | --- |
| Undiluted ThT | 0.41 | −0.53 | 0.079 | 0.395 | larger in Control |
| Diluted ThT | 0.12 | 0.14 | 0.901 | 0.901 | larger in LC |
| Pellet ThT | 0.64 | 0.88 | 0.728 | 0.901 | larger in LC |
| Pellet CD62P | −0.73 | −0.26 | 0.478 | 0.901 | larger in LC |
| Pellet Hoechst | −1.50 | −1.49 | 0.607 | 0.901 | larger in LC |

No experiment differs after correction (all *q* ≥ 0.40); undiluted ThT is the smallest (raw *p* = 0.079), again the only experiment carrying the size signal.

**Summary**

- **Abundance.** Long COVID elevates the amyloid-bearing (ThT, all three preparations) and activated-platelet (CD62P) fractions; the broader DNA-bearing, cell-derived pool (Hoechst) is unchanged (Table S1).
- **Dispersion.** The greater absolute spread in Long COVID is mean–variance coupling, not extra heterogeneity: log-scale Fligner–Killeen is non-significant for every experiment and log-scale SD ratios are near unity (Tables S2–S3).
- **Recovery.** ThT counts are not positively concordant across preparations; the one significant within-group association is a negative Diluted-vs-Pellet correlation in Long COVID (Table S4).
- **Composition.** The size distribution is largely preserved (equivalent for Diluted ThT, Pellet ThT and Hoechst from ±0.5 bin), with two exceptions: a suggestive shift toward smaller events in undiluted ThT (partly dispersion-driven) and markedly greater compositional heterogeneity of CD62P in Long COVID (Tables S5–S10). The phenotype is predominantly *more events of comparable size* rather than larger ones.
